# Benchmarking LLM-based Information Extraction Tools for Medical Documents

**DOI:** 10.64898/2026.01.19.26344287

**Authors:** Aaron Yu, Jochen Weile, Mélanie Courtot

**Author notes:** JW and MC contributed equally.

## Abstract

**Background:** Medical documents are a crucial resource for medical research around the world. While troves of valuable health data exist, they are largely computationally inaccessible as hard copies of unstructured text. Moreover, the persistent prevalence of fax machines in medical settings contributes to further degradation of document quality. Manual data extraction from these documents is time-consuming and resource intensive. However, large language models (LLMs) have recently shown great promise for automated digitization and information extraction (IE), greatly improving upon previous technologies in terms of accuracy.

**Objective:** Here we aim to survey the landscape of LLM-based IE tools with respect to their applicability to biomedical documents and to systematically benchmark them on synthetic documents with paired reference extractions. We put special focus on locally executable tools and models that suit privacy- and resource-constrained settings.

**Methods:** We reviewed LLM-based IE tools from the literature and assessed them with respect to their suitability for use in biomedical research. We found only one of these tools (NuExtract2) to satisfy our selection criteria and compared it to LLM foundation models prompted to perform extractions. We created 200 mock molecular test reports with paired reference data using a bespoke procedural generation approach and evaluated the tools’ performance across different prompting strategies, input modalities, and document qualities.

**Results:** We found model performance to be very sensitive to input modalities and quality. The best overall performance was observed for the proprietary GPT 4.1-mini, with an F_1_ score of 79.2% averaged across quality levels and modalities. The best performing open-weights model was Mistral-small 3.1 with an average F_1_ score of 76.2%. The NuExtract2 model was the only one able to run on regular laptop computers, but did not perform well. Surprisingly, we found the choice of one-shot over zero-shot prompts to only have a small effect size on extraction performance in most cases.

**Conclusions:** LLMs place the automatic processing of highly complex medical documents in resource-constrained and privacy-sensitive settings within reach, but still require human oversight.

**Availability:** Source code and data are available on GitHub [4] and on Zenodo [5], respectively.

## Introduction

### The critical role of early data digitization

The modern healthcare ecosystem faces a paradox: physicians and researchers are data-rich yet information-poor [23]. While vast quantities of invaluable clinical data are generated daily, a substantial portion remains locked away within paper-based records and non-standardized digital formats. Despite increased efforts toward digitization of resources, many healthcare institutions still rely on physical documents and analog technology such as photocopiers and fax machines to share information, thus adding distortions and artifacts to documents that decrease their legibility. This reliance on physical documents creates a critical bottleneck, rendering rich data sources largely inaccessible to computational analysis and modern health informatics.

As a consequence, medical information extraction (IE) and digitization are accomplished predominantly through manual curation. Researchers collect data from a wide variety of sources, including scanned documents, electronic health record (EHR)s, and faxed reports. This process is slow and labor-intensive, thus struggling to scale with the ever-increasing volume of medical data. Manual curation is also prone to human error, such as transcription mistakes or subjective interpretations, which can compromise data quality.

Automation of IE has been studied for some time. Early approaches rely on optical character recognition (OCR) followed by rule-based IE methods (e.g., regular expressions), but offer only a partial solution [29]. These systems are often brittle; they fail when encountering slight variations in document formatting and lack the semantic understanding required to interpret the context of clinical information accurately.

### Using large language models (LLMs) for data extraction

In recent years, transformer-based LLMs have emerged as state-of-the-art technologies capable of understanding and extracting structured information from unstructured documents with surprising accuracy [13, 2]. LLMs vastly improve the process of data extraction and processing thanks to their flexibility and greater accuracy.

The three primary types of transformer-based LLMs, Encoders, Decoders, and Encoder/Decoder hybrids, each offer unique advantages for clinical data processing tasks, positioning them as ideal candidates for addressing the complex extraction needs faced by researchers around Canada and the world. Encoder models such as the BERT family [7] create a latent semantic representation of text data, making them most commonly used for named entity recognition (NER) and IE tasks. Encoder/decoder hybrid models, such as BART [19], are often used in transformative sequence-to-sequence tasks such as translation or summarization. Pure decoder models such as OpenAI’s GPT, Mistral, and Google’s Gemini follow a sequence-to-sequence paradigm to generate new text based on a prompt, predicting what words are likely to follow [25, 21, 9]. Decoder models have attracted overwhelming attention due to their instruction-following capabilities and emergent zero-shot learning skills [30].

LLMs come in different sizes, typically expressed via the number of trained parameters. Larger models are usually more capable, but require increasingly expensive and power-hungry hardware to run and depend on larger amounts of training data. Therefore, the largest models are often proprietary and can only be accessed via the cloud. Much of the information fed into these commercial models is eventually used to train the next iteration of the model. While this may benefit future users with better performing technologies, it raises privacy issues.

Conversely, smaller models can be run locally without the requirement of sending data to third-party providers. However, to perform specialized task such as IE, smaller models often require fine-tuning, which depends on task-specific training data and machine-learning expertise.

Many published IE tools use the larger proprietary models via cloud solutions, due to their convenience. But in healthcare contexts, patient privacy and information security are paramount. In the absence of a trusted and certified cloud inference provider, locally hosted, open-source extraction tools are the only viable option for these applications.

Here, we study the feasibility of implementing LLMs for IE from biomedical documents. We benchmark a range of models and tools designed for extraction tasks. Specifically, we seek to answer the following questions: What local LLM-based options currently exist for biomedical data extraction? How do these tools perform in comparison to the cloud-based LLMs (which can be expected to be perform better, but are ineligible due to privacy issues)? What is the impact of common prompting strategies on extraction performance? Can multimodal tools and models capitalize on spatial context via image input compared to OCR-derived text input? And finally, which tool presents the best option for implementation into current workflows based on privacy, ease of use, and accuracy?

## Methods

### A survey of automated extraction tools

We reviewed the literature for LLM-based techniques and tools for IE tasks (Figure 1A). We used the following inclusion criteria for further investigation:

**Figure 1:**
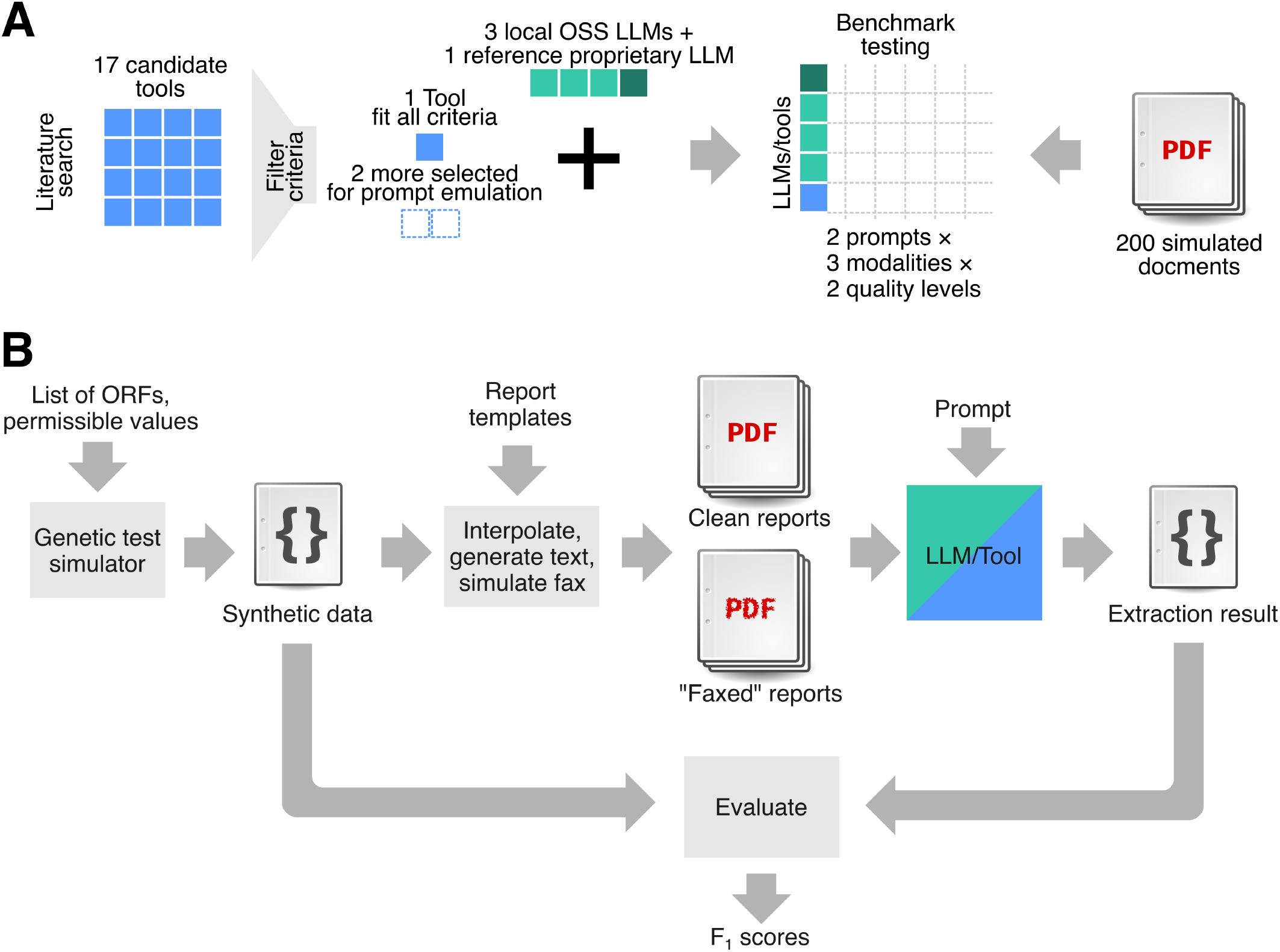
Overview of the benchmark process. A) We reviewed 17 LLM-based IE tools from the literature, finding only one (NuExtract2) to fit our selection criteria. We compared these tools against local open-weights foundation models, as well as one proprietary baseline model across different input modalities, input qualities and prompt designs on 200 synthetic molecular test reports. B) We implemented a generator tool for synthetic molecular test reports that generates possible test outcomes using a dataset of gene sequences. The generator creates tested gene panels as well as random variants within those genes. It then simulates the variants’ health impact. The simulated results are used to procedurally generate report texts which are interpolated into an array of different test report templates with different layouts and styles. A “fax”-distorted version of each document is also generated. The documents are fed into the tested model for data extraction and the result compared with the true information to calculate the model’s performance.

i. Tools should allow users to specify the desired entities to be extracted;
ii. Tools should function out-of-the-box and not require custom re-training or additional fine-tuning;
iii. Tools should be able to produce structured output, such as JavaScript Object Notation (JSON);
iv. Tools should have sufficient context size to enable processing full-text medical reports;
v. Tools should be open-source, such that code can be inspected for safety;
vi. Tools should either support general purpose data extraction, simple prompting of entity types to be extracted, or be purpose built for biomedical extraction tasks.

A survey of the available literature revealed 17 candidate tools for further examination. Of these, only one (NuExtract2) fully met our criteria. Supplementary Table S1 (Supplementary File 1) shows an overview of all candidates and which criteria they met.

NERRE [6] uses a fine-tuning approach with GPT3 [25] and Llama-2 [27]. It requires custom fine-tuning via a database of field-specific training examples. The model uses material science-specific extraction examples. Adaptation to biomedical use cases would thus require curation of applicable training data. Fine-tuning also requires the availability of a high-performance computing (HPC) environment, which may be inaccessible in resource constrained settings.

ATG [35] uses a constrained decoding approach to generate graph representations of extracted entities and their relationships. Unfortunately, model weights are not provided, requiring re-training of the model for new applications.

BioGPT [20] is a custom re-training of the GPT2 architecture on 15M PubMed abstracts. Unfortunately, the GPT2 architecture does not have a sufficiently large context window to support full-text documents. Preliminary tests showed poor IE performance, likely since GPT2 is a comparatively small decoder architecture intended for generative tasks.

GIX [11] is a tool for automatic literature search and molecular interaction extraction based on BioBERT and BERN2. Unfortunately, it does not generalize to extracting medical terms.

BERN2 [26] is an IE tool for molecular biology terms that also performs automatic entity normalization. Unfortunately, it currently only recognizes 9 biomedical entity types: Gene, disease, chemical, species, mutation, cell line, cell type, DNA, and RNA.

BioBERT [18] and Bio-LinkBERT [34] are versions of the BERT encoder architecture trained on biomedical texts. They can be used as foundations for many token-level tasks such as NER. However, they do require the design and training of task-specific post-processing networks.

GPT-NER [28] uses a few-shot prompting approach with examples automatically sampled from a vector database. This database of few-shot examples needs to be custom-tailored to the task domain. While GPT-NER does not work out of the box for biomedical extraction tasks we selected its prompt design for further manual testing.

Re-Rank-NER [32] uses an output re-ranking approach to improve the performance of a BART model for NER tasks. Unfortunately, the source code was not provided. The model would also require fine-tuning for biomedical document extraction.

UniversalNER [39] uses a targeted distillation approach to train custom small extraction models using larger teacher models. Unfortunately, it requires each entity type to be processed separately, which renders it not usable out of the box.

CancerBERT [38] is an encoder model trained on articles relating to cancer genomics. Like other encoder models, this would require the development and training of a post-processing neural network. In addition, neither the training data nor the model weights were provided.

P-ICL [16] uses few-shot prompts with a list of examples for each entity type to be extracted. Although the model weights were not made available, we selected the prompting strategy for further testing.

LTNER [33] uses a similar approach to GPT-NER. It uses contextualized markers to tag entities directly in the text sequence. Like GPT-NER, the tool also requires a vector database to retrieve relevant few-shot examples. While this made it difficult to fully replicate, we adapted the prompting approach for testing with fixed examples.

2INER [37] uses a fine-tuned model with few-shot prompts. The self-reported accuracy showed the tool to be approximately on-par with other NER or GPT-based methods. Unfortunately, no code or model weights were available for this tool.

NSSC [8] is a NER and entity normalization system for Spanish language clinical notes. As our test data was in English, it was unfortunately incompatible with our testing setup.

GliNER [36] is a bidirectional transformer encoder/decoder for any kind of entities to be extracted and labels text with entities. As a pure NER tool, it unfortunately does not allow users to specify fields to be extracted.

NuExtract2 [3] is a multi-modal model based on Qwen2.5-VL [1] that was fine-tuned for IE from unstructured documents. It can be installed via HuggingFace Transformers [31] and can be prompted for any extraction tasks. NuExtract2 met all our selection criteria and was therefore selected for further testing.

Notably, the P-ICL, LTNER and GPT-NER tools prominently featured few-shot prompting techniques. We combined these techniques into a few-shot prompt template to be tested across multiple foundation models in comparison with a zero-shot prompting approach (see Supplementary File 1 for prompt texts).

In addition to the selected purpose-built extraction tool, we also evaluated four general-purpose LLMs. Three of these are open-weights and can be run locally: Mistral-small 3.1 24B [21]; Llama 3.1 70B [27]; and Gemma 3 27B [10]. We installed these models via HuggingFace Transformers [31] and Ollama [22]. To serve as a baseline, we also tested a proprietary cloud-service LLM: GPT 4.1-mini [25].

### A synthetic benchmark dataset

To evaluate the selected models, we created a synthetic evaluation dataset simulating molecular test reports (Figure 1B). We started by creating JSON-formatted structured data in a three-stage process. First, we generated clinical metadata (such as test dates, lab information, clinic information, sample types, sequencing scopes, etc.) by randomly drawing from lists of allowed values. Secondly, we generated random gene panels from a set of 800 known disease genes. We then generated a set of genetic variants by drawing a Poisson-distributed random number of genes from the appropriate panel and simulating a random nucleotide change in each. We calculated the resulting amino acid changes, Human Genome Variation Society Nomenclature (HGVS) variant descriptors, associated database accessions, zygosity information, allele frequency and other variant-related information. In the final step, we simulated American College of Medical Genetics (ACMG) classifications via simulated evidence such as associated *in silico* variant impact predictions, functional assay outcomes, family history, etc.

Having created simulated structured data for genetic test outcomes, we then proceeded to generate simulated report documents for each dataset. This was achieved by interpolating the data into eleven different LaTeX templates corresponding to simulated testing sites. The templates were inspired by test report layouts from major testing sites and hospitals around Ontario, Canada, as well as online examples. Text sections such as full-text descriptions of the mutations and associated biological effects and interpretations in the documents were created synthetically from the generated data using procedural text generation. The interpolated LaTeX files were then compiled to PDF documents.

Finally, the documents were passed through an ImageMagick [15] script that simulates the effects of a fax machine or photocopier on image quality, by introducing small, random amounts of rotation, stretch and blur and applying Poisson noise. Example documents can be found in (Supplementary File 2). The R and LaTeX code used to generate the synthetic data and interpolated files can be found on GitHub [4].

In total, we generated 200 simulated clinical report datasets with reference data in JSON format and corresponding pairs of clean and image-distorted (“faxed”) PDF documents.

### Experimental setup and procedure

All models were run in a Python 3.14 virtual environment. We used a temperature parameter of *T* = 0 for all LLM calls. This parameter controls the distribution of probability density across output tokens. At *T* = 0 the token with the highest score receives 100% of the probability density and is thus always sampled. This serves to force consistent output, restricting the LLM’s creativity and constraining the model to the most likely tokens. The Mistral-small 3.1 installation for Ollama ships with a default setting that restricts its context window. To override this restriction, we set the num_ctx parameter to 128, 000 such as to unlock its full context size. All tested models, including the large LLama 3.1 70B were hosted on a HPC platform. NuExtract2 was also run on a standard MacBook Pro with 24GB of RAM and an M4 chip.

In the first step, images of each page were extracted using Poppler [14] and then parsed into text via EasyOCR [17]. For multimodal models (featuring both vision and text input capabilities) images were directly supplied to them alongside the prompts. As a baseline control, models were also tested on the original raw document texts in Markdown format, which were compiled directly from the interpolated LaTeX sources using Pandoc.

The LLMs were tested with two types of IE prompts: A zero-shot prompt (providing no examples to the LLM) inspired by the prompt used in P-ICL [16] and a oneshot prompt inspired by those used in LTNER [33] and GPT-NER [28]. Each prompt instructed the respective model to return its output in JSON format for easy comparison. The zero-shot prompt comprised a list of data fields to extract and, where appropriate, a list of allowed terms for each field, together with a blank JSON template for the output. The one-shot prompt contained the same elements with the addition of one text example and its expected output. See Supplementary File 1 for prompt texts.

The prompts were provided to the models alongside the raw text, OCR-processed text, or raw images respectively. The NuExtract2 model required a different input format: (1) a specific JSON template that included field names, their expected type, and options within that field; (2) a separate input file for any one-shot examples; and (3) a concatenation of the page images into a single image.

Finally, the extraction results were compared to the reference data. A detailed description of the scoring procedure, including multiple examples, can be found in the Supplementary Methods (Supplementary File 1). Briefly, the relevant reference data for each report was trimmed down to the subset of fields that were compatible with the respective document template. Next, the extracted data was compared value by value for matches. Most fields required perfect matches to count as correct. However, for certain fields containing more text-like data (e.g. hospital names, testing context), partial matches were allowed. This was calculated using character-level alignments via a simplified Needleman-Wunsch algorithm [24]. This scoring approach strikes a balance between punishing technically incorrect extractions and rewarding partial correctness. Based on these full or partial matches, we then counted the number of true positives (TPs) (correct matches), false positives (FPs) (surplus or hallucinated extractions) and false negatives (FNs) (missing extractions) in each field.

We then calculated the precision, recall and F_1_ score for each JSON output (with precision defined as the fraction of TPs out of all extracted entities (i.e. TP+FP) and recall defined as the fraction of TPs out of all entities that were meant to be extracted (i.e. TP+FN), and F_1_ score defined as the harmonic mean of precision and recall). The F_1_ score was chosen as the primary metric of comparison as it provides a balanced measure of a model’s performance.

We also tracked whether outputs adhered to the JSON formatting requirement at 5 different quality levels: (1) Output only consists of valid JSON; (2) valid JSON embedded in a Markdown code block; (3) valid JSON buried in the middle of text, detectable via searching for pairs of matching braces; (4) Invalid yet repairable JSON (via removal of comments and fixing of missing or superfluous commas, brackets or braces) either buried or embedded in text as above; and (5) Unrepairable, unparseable output. Unparseable outputs were assigned precision, recall and F_1_ scores of 0, respectively.

## Results

Using our testing framework, we evaluated all five models on our benchmark of 200 simulated molecular test report documents across two different prompting strategies, two input modalities and two quality levels plus the raw text input baseline. Four out of five models had multimodal input capabilities and were tested on all inputs. Llama 3.1 was only able to accept text inputs. GPT 4.1-mini was accessed via a cloud API, NuExtract2 was run locally on a laptop computer via HuggingFace Transformers, and the remaining tools and models were open-weights and were run locally on an HPC cluster via an Ollama server. Outputs were compared to their references, F_1_ scores were calculated and averaged across all outputs per hyperparameter combination.

First, we examined how well tool output conformed to the required JSON format. Most models provided Mark-down-embedded JSON. Only LLama 3.1 and NuExtract provided pure perfect JSON for a substantial number of tests. Gemma 3, Llama 3.1 and Mistral-small 3.1 produced small amounts of lower quality outputs, where JSON was buried in text or required repair (22/2,000 = 1.1% ; 13/1,200 = 1.1%; and 23/2,000 = 3.1%, respectively). NuExtract2 behaved unexpectedly. Image and OCR inputs for this model largely resulted in perfect JSON. However, when given the original raw text in Markdown format, NuExtract2 overwhelmingly responded with unusable output. This behavior turned out to be caused by its built-in input processing function, which combines the prompt, JSON-template, one-shot examples and text input into a fixed Markdown structure, such that providing Markdown-formatted input confused the model’s ability to determine where the fixed Markdown template ends and the document starts (Figure 2A).

**Figure 2:**
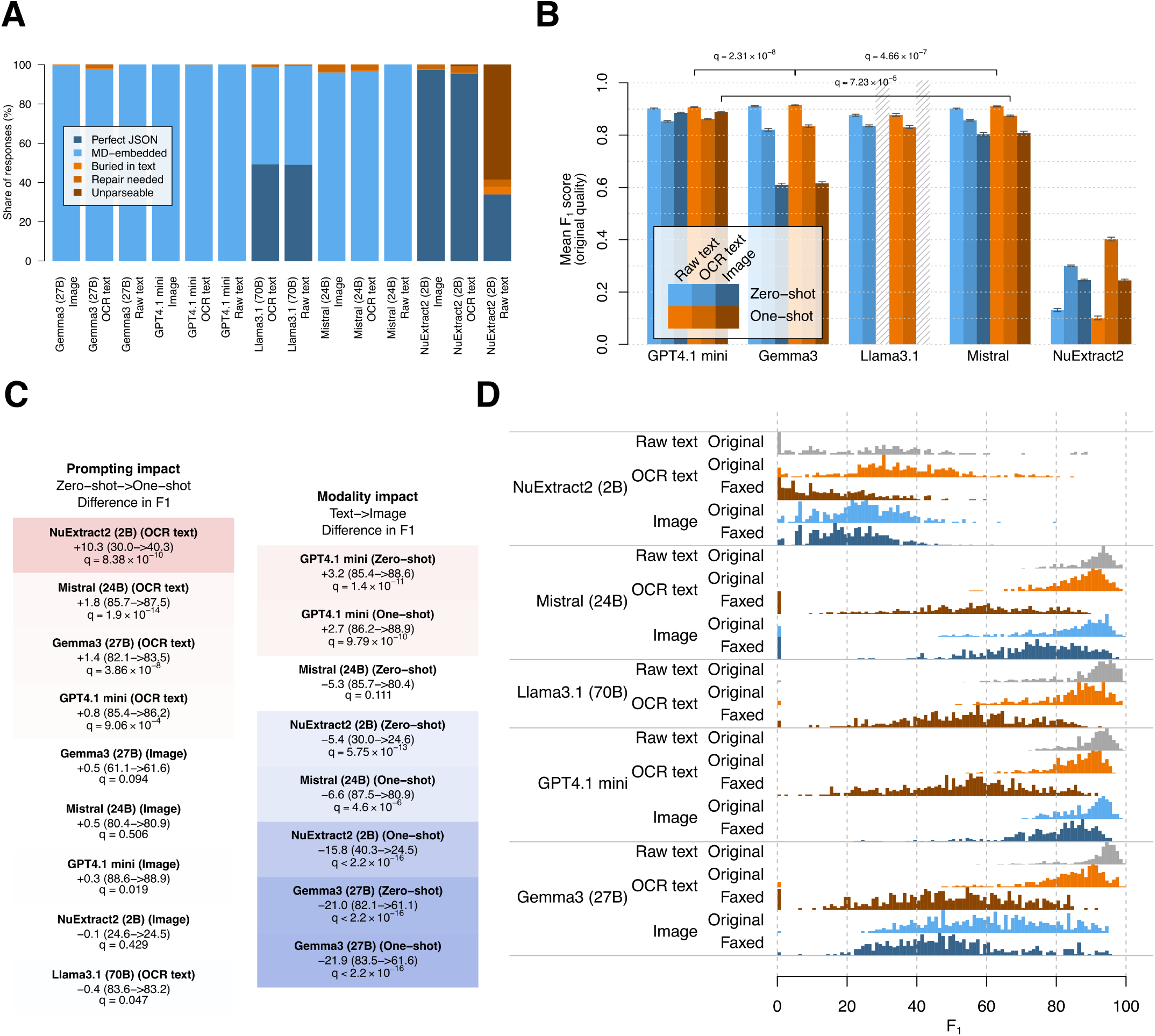
Benchmark results for the tested models. A) The JSON parseability grade for each tested tool. See Methods for full explanation of each grade. (B) Mean F_1_ scores of models given undistorted inputs split by prompt and input modality. Error bars indicate standard error. Brackets indicate pWSRT, with FDR-corrected q-values. (C) Differences in mean F_1_ between zero-shot and one-shot prompts (left) and OCR-derived text and direct image input, respectively, for each applicable tool. q-values are given for FDR-corrected pWSRT tests. (D) Distributions of F_1_ scores for tested models using zero-shot prompts, split by input modality (image vs OCR-derived text) and input quality (clean PDF vs fax-simulated).

We then compared the mean extraction F_1_ scores of different combinations of models, prompts and input modalities (Figure 2B, Supplementary Figure S1). As expected, the GPT 4.1-mini baseline model performed consistently well across all categories, at an average of 79.2% F1. On raw document text, GPT 4.1-mini reached a peak performance of 90.7% F_1_ (using one-shot prompts). The best performing open-weights models in this category were Gemma 3 and Mistral-small 3.1, reaching peaks of 91.6% and 91.1% F_1_ respectively. While statistically significant (*q* = 2.31 · 10^*−*8^ and *q* = 4.66 · 10^*−*7^ False discovery rate (FDR)-corrected paired Wilcoxon signed-rank test (pWSRT), respectively), the differences are very small. Their average performances across all quality levels and input modalities differed more markedly however, with Gemma 3 at 66.7% and Mistral-small 3.1 at 76.2% F1.

Given high quality PDFs (ingestable either as OCR-derived text or direct image input), GPT 4.1-mini led similarly with 88.9% F_1_ (using one-shot prompts and image inputs), followed by Mistral-small 3.1 at 87.5% F_1_ (using one-shot prompts and EasyOCR) with a similarly minor difference of 1.4 percentage points (*q* = 7.23·10^*−*5^, FDR-corrected pWSRT)

The distortions caused by the simulated fax machine had a strong effect on model performances across the board. Generally, direct image inputs were superior to external OCR in this scenario. The best performance was once again seen for GPT 4.1-mini at 78.4% F_1_ (given oneshot prompts and direct image inputs). Here, the best open-weights model was once again Mistral-small 3.1 at 70.8% F_1_ (given zero-shot and direct image input); with a *−* 7.6 percentage point difference (*q* = 9.57 · 10^*−*15^, FDR-corrected pWSRT).

The NuExtract2 model performed poorly in all scenarios compared to the bigger models. While this could be expected due to its much smaller size at only 2B parameters, it is nonetheless disappointing given the prospect of its minimal hardware requirements.

The use of one-shot instead of zero-shot prompts showed only very minor effects in most scenarios. NuExtract2 was the only model to see substantial performance increases (+10.3 F_1_ percentage points) given one-shot prompts (*q* = 8.38 · 10^*−*10^, FDR-corrected pWSRT). While changes for Mistral-small 3.1, Gemma 3, and GPT 4.1 mini were also statistically significant, the effect sizes were numerically negligible (see Figure 2C for q-values and effect sizes).

Direct image input generally appeared to perform worse than external OCR for most models (Figure 2C). GPT 4.1-mini stood out as the notable exception, seeing a small +3.2 and +2.7 percentage point increase (*q* = 1.4 · 10^*−*11^ and *q* = 9.79 · 10^*−*10^; FDR-corrected pWSRT) for zero- and one-shot prompts, respectively. The greatest impact was seen for Gemma 3, showing a 21.0 and 21.9 percentage point F_1_ difference given zero-shot and one-shot prompts, respectively.

However as mentioned above, the impact of input modality was highly dependent on document quality. When given fax-distorted documents, direct image inputs performed better than EasyOCR for all vision models, most noticeably so for GPT 4.1-mini (+24.9 and +24.4 percentage points for zero- and one-shot, respectively) and Mistral-small 3.1 (+19.6 and +16.7 points for zero- and one-shot, respectively).

When examining the shapes of the F_1_ score distributions, more nuance emerges (Figure 2D). Fax distorted inputs do not merely shift the mean of the F_1_ distribution, but greatly increase the variance of the outputs, often reaching from below 20% F_1_ to above 90% F1. Similarly, the use of image inputs in well performing models often adds a long tail to score distributions.

We next wondered whether the different layout templates had an impact on model performance. The templates differ in format, presentation and the volume of information contained. Unsurprisingly, the synthetic templates J and K were close to the extreme ends of the observed difficulty levels, as they were designed to cover a very minimal and maximal amount of extractable fields, respectively. Template H appeared to be generally easier to process as well due to its relatively simple layout and minimal use of tables. This primarily helped OCR and direct image inputs. Conversely, template C was among the most difficult to process. It includes large redactions and contains many visually complex tables. Another outlier was seen for template G, as it emulates a single-gene test result instead of a gene panel, thus requiring only a single gene name to be extracted and thus providing less opportunity for extraction. (Figure 3).

**Figure 3:**
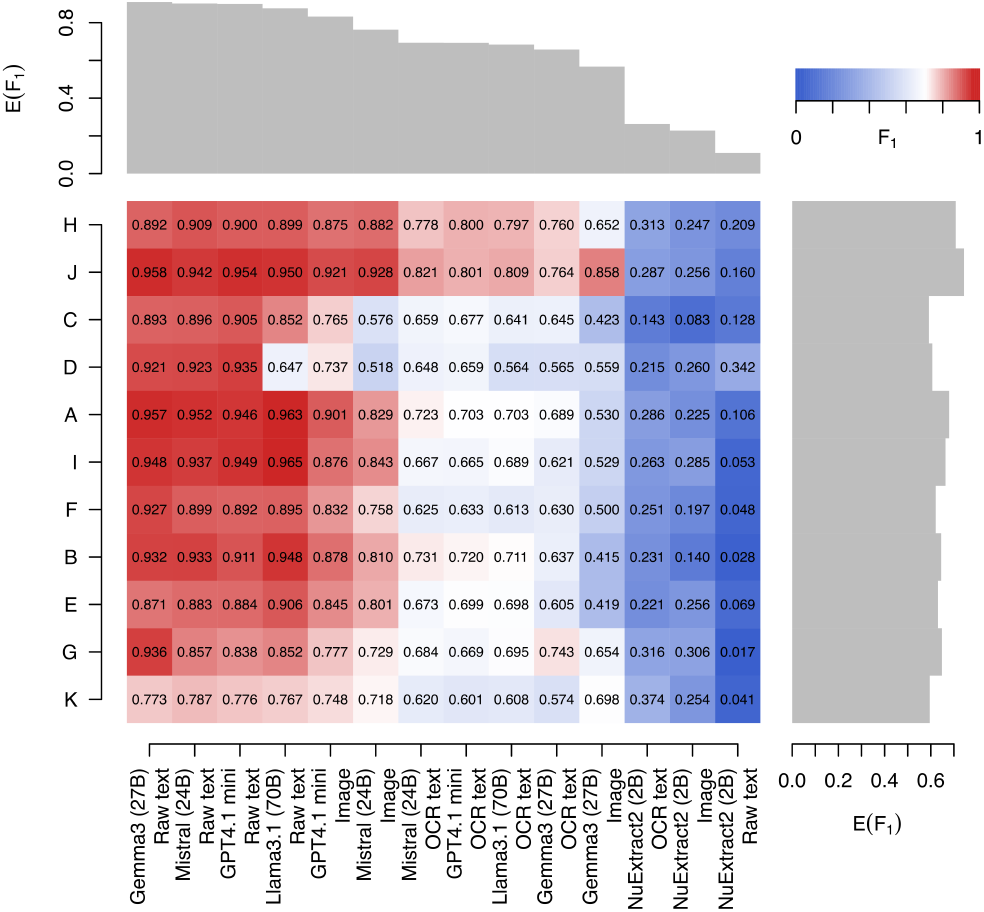
A heatmap comparing the mean F_1_ score for combinations of specific report layout templates and the model/input modality combinations.

Next, we wondered what types of information were most difficult to extract across models. We therefore examined precision and recall for each extractable field type separately (Supplementary Figure S2). Fields like the report type, analysis type and testing context, which often needed to be contextually inferred, unsurprisingly suffered the lowest recall.

For the NuExtract model, genetic information, namely gene symbols, transcript IDs, and variants appeared to be the most challenging, suffering the lowest extraction precision. In most layout templates, gene information was densely packed within tables which required models to take into account the spatial positioning of elements to interpret their relationships to each other. This may have contributed to the lower success rate on this type of data.

In addition, gene symbols and transcript IDs appear as arbitrary combinations of letters and numbers, which makes them hard for OCR to recognize and for LLMs to tokenize and process. Depending on the specific typeface used in a given report layout, characters like 0/O, 1/I and S/5 can be difficult to distinguish when not in context of natural language words. As such, a gene symbol like PIP5L1 might be mistakenly read as P1PSLI.

Surprisingly, for all other models, only very few errors related to detailed variant information. This was likely due to the prominent formatting and clarity of sections in the clinical reports related to variant location, prevalence, and related genetic information, as variant information is the primary topic of the reports.

## Discussion

Here we have evaluated the performance of LLMs tools for IE from synthetic medical documents using simulated molecular testing reports. While results are based on synthetic documents, they may help guide the use of such tools as part of digitization and data extraction tasks for biomedical research. Below, we discuss insights gained from the effects of document qualities, modalities and prompts, before discussing our conclusions on the suitability of the tested models for different purposes.

Our results showed that the design of document layouts can have a meaningful impact on interpretability by extraction tools. Depending on their internal document designs, individual clinical locations may thus benefit differently from such tools. This emphasizes the importance of choosing large, legible typefaces and clear layouts for clinical documents not only for accessibility reasons, but also for improving automated extraction performance.

Beyond document complexity, the visual quality of the document also appears to have a major impact on extraction success. Image distortions introduced by the fax-machine simulator add to the stack of potential errors: they compound OCR failures, which in turn compound context misinterpretations by the LLM, which in turn compound the ability to extract the needed information and output it in the desired structured format. This is especially true for codes and symbols such as gene names, which contain numerals and letters prone to OCR misinterpretation (e.g. confusing “0” with “O” or “1” with “I”). This often affected extraction in gene information, especially gene names. However, models with advanced contextual reasoning abilities, notably GPT 4.1-mini, can be expected to be better at rectifying such OCR-induced mistakes during extraction, for example being able to distinguish that BRCAl should actually be BRCA1. Accordingly, documents that underwent our simulated fax-machine procedure saw an average F_1_ degradation of 30.0 percentage points when relying on external OCR and 9.2 points when using direct image inputs.

Document quality also had an impact on performance variability. While most models performed relatively consistently on original-quality documents (ranging approximately between 80% and 100% F1), fax-distorted documents yielded wildly varying scores (ranging between approximately 10% and 90%, Figure 2D).

We were surprised to see that using one-shot prompts over zero-shot prompts did not greatly improve extraction results. However, it is conceivable that document complexity may have played a role here as well. Other prompting strategies may be worth exploring in future research.

Prompt design can nonetheless have major impact on extraction performance. An earlier version of this study had accidentally included mistakes in the prompts that while appearing minor to a human eye (such as missing commas in the JSON format description) led to major performance degradations. A detailed ablation study of the effects of such mistakes would be useful to consider in the future.

### Model suitability

From a practical perspective, the most reliable model was the GPT 4.1-mini baseline. It consistently ranked among the best results, especially when provided fax-distorted image inputs. However, as a proprietary model only accessible via remote API, it is not suitable for privacy-sensitive applications such as the processing of medical documents.

Given these caveats, the Mistral-small 3.1 model appears to offer the best combination of F_1_ performance (91.1% on raw text, 87.5% on clean PDFs, and 70.7% on distorted PDFs), speed and local deployability. However, human oversight is still necessary, especially when working with complex, low-quality documents.

The NuExtract2 model unfortunately came in last. Given its small size and ability to run on laptop hardware, it may still be suitable for simpler extraction tasks under strict hardware constraints. Its apparent incompatibility with Markdown input, as reflected in its poor performance on the raw document texts compared to OCR or images, was disappointing. The additional requirement of concatenating pages into single images further complicates its usage. A larger 4B version of NuExtract2 exists, which also runs on laptop hardware. We tested it on a MacBook Pro with 24GB of RAM and an M4 chip as well as on a MacBook with 16GB of RAM and an M1 chip. In preliminary tests the 4B performed better on OCR-text than its 2B counterpart, but we were unable to make it accept image inputs.

The Gemma 3 model performed admirably. However, its vision capabilities turned out to be disappointing, lagging far behind using EasyOCR input.

The Llama model performed generally in the same ballpark as Gemma and Mistral. As a text-only model it is fully dependent on the quality of the upstream OCR tool (in this case EasyOCR).

Although error rates are impressively low, even top performing models would currently require manual verification steps before deployment in biomedical research settings. While human verification is resource- and time-intensive, it would likely still present an improvement over fully manual extraction. Verification would be a necessary step to mitigate risks associated with erroneous data extractions, thereby maintaining the data integrity critical for research.

### Infrastructure and accessibility

Steep hardware requirements for using frontier-class open-weights LLMs in data extraction for biomedical research do little to alleviate the equity issues faced by rural and remote communities. However, using such large models may not be necessary. The smaller local models tested here can be executed on a laptop computer. Quantized versions of Gemma 3 (27B) and Mistral-small 3.1 (24B) can theoretically run on a high-end MacBook Pro (albeit monopolizing a majority of its memory). The NuExtract2 model stands out for its very small footprint, although its performance was disappointing. An alternative solution would be the use of remote services. As discussed above, proprietary model APIs such as the one we used for GPT 4.1-mini here are incompatible with patient privacy requirements. However, they should not be the only option. A certified end-to-end encrypted biomedical data extraction service could fill an important niche here.

### Limitations

Our benchmark results have several limitations. First, the artificial nature of the test dataset and the limited variety of document types, formats and layouts tested might constrain the generalizability of these results. While we used a variety of eleven different layouts and structures inspired by various clinical sites across Ontario and online examples, our synthetic dataset may not fully reflect real-world scenarios and the potentially broader scope of genomic information and types of clinical reports.

We aimed to offset this limitation by introducing distortions to our synthetic documents that mimic the effects of fax or photo copier machines. The distortions appear visually realistic, but machine vision does not necessarily rely on the same features as human vision. Real world documents can also suffer from other defects that we did not cover here, such as added handwritten annotations, highlights, coffee stains, or wrinkles.

Second, we excluded many tools from our tests for requiring more complex setup. One such requirement was the use of vector databases to enable retrieval-augmented generation (RAG) workflows and few-shot example selection. Such approaches may improve accuracy and reduce the time required for extraction by enabling models to first localize relevant information within lengthy clinical reports before performing extraction. However, this requires a dedicated dataset with partitioned examples. While we consider these improvements out of scope for this benchmark, future research may incorporate these ideas.

Third, we only examined two broad prompting strategies. However, a wide range of other strategies exists, featuring a great amount of nuance which we cannot capture here. Even though our results only showed limited differences between zero-shot and one-shot prompts, other approaches may impact model performance more drastically. Future research may include additional prompting strategies. Having seen incidental evidence of performance degradation due to typos and formatting errors in prompts during experimentation furthermore suggests that a dedicated ablation study on the effects of prompt formats, formulations and errors may be warranted.

Fourth, we only tested LLM-based tools in this benchmark, forgoing older rule-based IE systems. However, we did not find any such systems conforming to our selection criteria. Most rule-based extraction systems are bespoke to specific tasks and domains and often focus on entity-relationship extractions.

Finally, due to our exclusion criteria, we only tested five different models. The LLM field is currently very active with new models being published every month. Further studies could expand to benchmarking a wider range of foundation models such as Qwen [1] or DeepSeek [12].

### Future work

We may expand our benchmark in the future to include newer models. Future research may also explore more advanced architectures such as RAG to enhance extraction accuracy and efficiency. It would also be useful to expand our benchmark beyond molecular test report documents towards other types of medical documents. Finally, it would be worth exploring to what extent task-specific fine-tuning of the larger multimodal models such as Gemma 3 can improve their extraction performance.

### Conclusion

This research aimed to review and benchmark LLM-based IE tools from the current literature on complex clinical documents, namely molecular test reports. We tested five different models and while their accuracy was impressive, they still require human oversight and manual validation. This underlines the need for further research into improving these tools.

## Data Availability

All data and code produced in the present study are available on GitHub and Zenodo.

https://github.com/courtotlab/extraction-benchmark

https://doi.org/10.5281/zenodo.22904341

## AI disclosure

The authors declare that generative AI was not used in the writing of this manuscript text. However, for the underlying experimental pipeline code, GitHub Copilot was used to assist with coding some portions of R and python scripts for the document mockup generator and scoring functions.

## Data availability

Source code and data are available on GitHub [4] and on Zenodo [5], respectively.

## Author contributions

J.W., A.Y. and M.C. conceived of the experiments, A.Y. conducted the experiments, J.W., A.Y. analyzed the results. A.Y. and J.W. wrote the manuscript with edits from M.C. J.W. and M.C. supervised the project.

## Acronyms

ACMG: American College of Medical Genetics classification standard: A set of rules governing which types of evidence contribute in which way to classify genetic variants with respect to their clinical implications, ranging from Benign via Variants of Uncertain Significance (VUS) to Pathogenic. 4
EHR: Electronic health record: A comprehensive digital medical history of a patient accessible and maintainable by multiple healthcare providers. 2
FDR: False discovery rate correction: A method by Benjamini and Hochberg to adjust the p-values of statistical tests controlling for the underlying false discovery rate.. 6
FN: False negatives: The number of items that should have been extracted but were missed. 5
FP: False positives: The number of incorrectly extracted items. 5
HGVS: Human Genome Variation Society Nomenclature: Official syntax for describing variants in the human genome, governs descriptions of variants at the genomic, coding sequence, transcript, and protein level. 4
HPC: High-performance computing: The use of large amounts of aggregated powerful computing resources. 3, 5
IE: Information Extraction: The task of extracting structured data or information from unstructured text. 1, 2, 4, 5, 8, 9
JSON: JavaScript Object Notation: A data serialization format expressing objects as embedded combinations of dictionaries and lists. 2, 4, 5, 6, 8
LLM: Large language model: A pre-trained transformer model that models natural language. 1, 2, 4, 5, 7, 8, 9
NER: Named entity recognition: The task of identifying specific named entities in natural language text. 2, 4
OCR: Optical Character Recognition: A machine learning method that recognizes text in images and outputs that text. 2, 5, 6, 7, 8, 9
pWSRT: paired Wilcoxon signed-rank test: A non-parametric statistical test comparing the locations of two populations using paired samples.. 6
RAG: Retrieval-augmented generation: A technique for providing relevant knowledge to an LLM for a given query via a neural retriever. 9
TP: True positives: The number of correctly extracted items. 5

## References

[1] S. Bai, K. Chen, X. Liu, J. Wang, W. Ge, S. Song, K. Dang, P. Wang, S. Wang, J. Tang, H. Zhong, Y. Zhu, M. Yang, Z. Li, J. Wan, P. Wang, W. Ding, Z. Fu, Y. Xu, J. Ye, X. Zhang, T. Xie, Z. Cheng, H. Zhang, Z. Yang, H. Xu, and J. Lin. Qwen2.5-vl technical report, 2025. URL https://arxiv.org/abs/2502.13923.

[2] D. Chen, S. Alnassar, K. Avison, R. Huang, and S. Raman. Large language model applications for health information extraction in oncology: Scoping review. JMIR Cancer, 11, 2025. ISSN e65984. doi: 10.2196/65984. URL https://cancer.jmir.org/2025/1/e65984.

[3] A. Constantin, L. Cripwell, N. Fradet, S. Dréano, and E. Bernard. Nuextract: A foundation model for structured extraction, June 2024. URL https://github.com/numindai/nuextract. original-date: 2025-02-18T14:07:59Z.

[4] CourtotLab. Extraction benchmark GitHub repository, 2026. URL https://github.com/courtotlab/extraction-benchmark.

[5] CourtotLab. Information extraction benchmark Zenodo repository, 2026. URL 10.5281/zenodo.22904341.

[6] J. Dagdelen, A. Dunn, S. Lee, N. Walker, A. S. Rosen, G. Ceder, K. A. Persson, and A. Jain. Structured information extraction from scientific text with large language models. Nature Communications, 15(1):1–14, Feb. 2024. ISSN 2041-1723. doi: 10.1038/s41467-024-45563-x. URL https://www.nature.com/articles/s41467-024-45563-x.

[7] J. Devlin, M.-W. Chang, K. Lee, and K. Toutanova. BERT: Pre-training of deep bidirectional transformers for language understanding. In Proceedings of the 2019 Conference of the North American Chapter of the Association for Computational Linguistics: Human Language Technologies, volume 1 (Long and Short Papers), pages 4171–4186, Stroudsburg, PA, USA, 2019. Association for Computational Linguistics. doi: 10.18653/v1/N19-1423.

[8] Á. García-Barragán, A. Sakor, M.-E. Vidal, E. Menasalvas, J. C. S. Gonzalez, M. Provencio, and V. Robles. NSSC: a neuro-symbolic AI system for enhancing accuracy of named entity recognition and linking from oncologic clinical notes. Medical & Biological Engineering & Computing, 63(3):749–772, 2025. ISSN 0140-0118. doi: 10.1007/s11517-024-03227-4. URL https://www.ncbi.nlm.nih.gov/pmc/articles/PMC11891111/.

[9] Gemini Team. Gemini: A family of highly capable multimodal models. arXiv [cs.CL], page 2312.11805, Dec. 2023. doi: 10.48550/arXiv.2312.11805.

[10] Gemma Team. Gemma 3 technical report, 2025. URL https://arxiv.org/abs/2503.19786.

[11] J. K. Gill, M. Chetty, S. Lim, and J. Hallinan. Large language model based framework for automated extraction of genetic interactions from unstructured data. PLOS ONE, 19(5):e0303231, May 2024. ISSN 1932-6203. doi: 10.1371/journal.pone.0303231. URL https://journals.plos.org/plosone/article?id=10.1371/journal.pone.0303231.

[12] D. Guo, D. Yang, H. Zhang, J. Song, P. Wang, Q. Zhu, R. Xu, R. Zhang, S. Ma, X. Bi, X. Zhang, X. Yu, Y. Wu, Z. F. Wu, Z. Gou, Z. Shao, Z. Li, Z. Gao, A. Liu, B. Xue, B. Wang, B. Wu, B. Feng, C. Lu, C. Zhao, C. Deng, C. Ruan, D. Dai, D. Chen, D. Ji, E. Li, F. Lin, F. Dai, F. Luo, G. Hao, G. Chen, G. Li, H. Zhang, H. Xu, H. Ding, H. Gao, H. Qu, H. Li, J. Guo, J. Li, J. Chen, J. Yuan, J. Tu, J. Qiu, J. Li, J. L. Cai, J. Ni, J. Liang, J. Chen, K. Dong, K. Hu, K. You, K. Gao, K. Guan, K. Huang, K. Yu, L. Wang, L. Zhang, L. Zhao, L. Wang, Zhang, L. Xu, L. Xia, M. Zhang, M. Zhang, Tang, M. Zhou, M. Li, M. Wang, M. Li, N. Tian, P. Huang, P. Zhang, Q. Wang, Q. Chen, Q. Du, Ge, R. Zhang, R. Pan, R. Wang, R. J. Chen, R. L. Jin, R. Chen, S. Lu, S. Zhou, S. Chen, S. Ye, S. Wang, Yu, S. Zhou, S. Pan, S. S. Li, S. Zhou, S. Wu, Yun, T. Pei, T. Sun, T. Wang, W. Zeng, W. Liu, W. Liang, W. Gao, W. Yu, W. Zhang, W. L. Xiao, W. An, X. Liu, X. Wang, X. Chen, X. Nie, X. Cheng, X. Liu, X. Xie, X. Liu, X. Yang, X. Li, X. Su, X. Lin, X. Q. Li, X. Jin, X. Shen, X. Chen, X. Sun, X. Wang, X. Song, X. Zhou, X. Wang, X. Shan, Y. K. Li, Y. Q. Wang, Y. X. Wei, Y. Zhang, Y. Xu, Y. Li, Y. Zhao, Y. Sun, Y. Wang, Y. Yu, Y. Zhang, Y. Shi, Y. Xiong, Y. He, Y. Piao, Y. Wang, Y. Tan, Y. Ma, Y. Liu, Y. Guo, Y. Ou, Y. Wang, Y. Gong, Y. Zou, Y. He, Y. Xiong, Y. Luo, Y. You, Y. Liu, Y. Zhou, Y. X. Zhu, Y. Huang, Y. Li, Y. Zheng, Y. Zhu, Y. Ma, Y. Tang, Y. Zha, Y. Yan, Z. Z. Ren, Z. Ren, Z. Sha, Z. Fu, Z. Xu, Z. Xie, Z. Zhang, Z. Hao, Z. Ma, Z. Yan, Z. Wu, Z. Gu, Z. Zhu, Z. Liu, Z. Li, Z. Xie, Z. Song, Z. Pan, Z. Huang, Z. Xu, Z. Zhang, and Z. Zhang. DeepSeek-R1 incentivizes reasoning in llms through reinforcement learning. Nature, 645(8081):633–638, Sept. 2025. ISSN 1476-4687. doi: 10.1038/s41586-025-09422-z. URL http://dx.doi.org/10.1038/s41586-025-09422-z.

[13] K. He, R. Mao, Q. Lin, Y. Ruan, X. Lan, M. Feng, and E. Cambria. A survey of large language models for healthcare: from data, technology, and applications to accountability and ethics. Inf. Fusion, 118:102963, Jan. 2025. doi: 10.1016/j.inffus.2025.102963.

[14] K. Høgsberg, A. Astals Cid, C. Garcia Campos, A. Johnson, and P. Toscano. Poppler the pdf rendering library, 2005. URL https://gitlab.freedesktop.org/poppler/poppler.

[15] ImageMagick Studio LLC. Imagemagick. URL https://imagemagick.org.

[16] G. Jiang, Z. Ding, Y. Shi, and D. Yang. P-ICL: Point in-context learning for named entity recognition with large language models. ArXiv, abs/2405.04960, May 2024. doi: 10.48550/arXiv.2405.04960.

[17] R. Kittinaradorn. EasyOCR: Ready-to-use OCR with 80+ supported languages and all popular writing scripts including latin, chinese, arabic, devanagari, cyrillic and etc, June 2020. URL https://github.com/JaidedAI/EasyOCR.

[18] J. Lee, W. Yoon, S. Kim, D. Kim, S. Kim, C. H. So, and J. Kang. BioBERT: a pre-trained biomedical language representation model for biomedical text mining. Bioinformatics, 36(4):1234–1240, Feb. 2020. ISSN 1367-4803. doi: 10.1093/bioinformatics/btz682. URL https://doi.org/10.1093/bioinformatics/btz682.

[19] M. Lewis, Y. Liu, N. Goyal, M. Ghazvininejad, A. Mohamed, O. Levy, V. Stoyanov, and L. Zettlemoyer. BART: Denoising sequence-to-sequence pretraining for natural language generation, translation, and comprehension. In Proceedings of the 58th Annual Meeting of the Association for Computational Linguistics, pages 7871–7880, Stroudsburg, PA, USA, 2020. Association for Computational Linguistics. doi: 10.18653/v1/2020.acl-main.703.

[20] R. Luo, L. Sun, Y. Xia, T. Qin, S. Zhang, H. Poon, and T.-Y. Liu. BioGPT: generative pre-trained transformer for biomedical text generation and mining. Briefings in Bioinformatics, 23(6):bbac409, Nov. 2022. ISSN 1477-4054. doi: 10.1093/bib/bbac409. URL https://doi.org/10.1093/bib/bbac409.

[21] Mistral-AI. Magistral. arXiv [cs.CL], page 2506.10910, June 2025. doi: 10.48550/arXiv.2506.10910.

[22] J. Morgan, M. Yang, B. MacDonald, D. Hiltgen, and M. Williams. Ollama: Get up and running with llama 3.3, DeepSeek-R1, Phi-4, Gemma 3, Mistral Small 3.1 and other large language models, June 2023. URL https://github.com/ollama/ollama.

[23] T. B. Murdoch and A. S. Detsky. The inevitable application of big data to health care. JAMA, 309 (13):1351, Apr. 2013. doi: 10.1001/jama.2013.393.

[24] S. Needleman and C. Wunsch. A general method applicable to the search for similarities in the amino acid sequence of two proteins. J Mol Biol., 48(3):443–453, 1970. doi: 10.1016/0022-2836(70)90057-4.

[25] OpenAI. GPT-4 technical report. arXiv [cs.CL], page 2303.08774, Mar. 2023. doi: 10.48550/arXiv.2303.08774.

[26] M. Sung, M. Jeong, Y. Choi, D. Kim, J. Lee, and J. Kang. BERN2: an advanced neural biomedical named entity recognition and normalization tool. Bioinformatics, 38(20):4837–4839, Sept. 2022. ISSN 1367-4803. doi: 10.1093/bioinformatics/btac598. URL https://www.ncbi.nlm.nih.gov/pmc/articles/PMC9563680/.

[27] H. Touvron, T. Lavril, G. Izacard, X. Martinet, M.-A. Lachaux, T. Lacroix, B. Rozière, N. Goyal, E. Hambro, F. Azhar, A. Rodriguez, A. Joulin, E. Grave, and G. Lample. LLaMA: Open and Efficient Foundation Language Models, Feb. 2023. URL https://arxiv.org/abs/2302.13971. ADS Bibcode: 2023arXiv230213971T.

[28] S. Wang, X. Sun, X. Li, R. Ouyang, F. Wu, T. Zhang, J. Li, G. Wang, and C. Guo. GPT-NER: Named entity recognition via large language models. In Findings of the Association for Computational Linguistics: NAACL 2025, pages 4257–4275, Stroudsburg, PA, USA, 2025. Association for Computational Linguistics. doi: 10.18653/v1/2025.findings-naacl.239.

[29] Y. Wang, L. Wang, M. Rastegar-Mojarad, S. Moon, F. Shen, N. Afzal, S. Liu, Y. Zeng, S. Mehrabi, S. Sohn, and H. Liu. Clinical information extraction applications: A literature review. J. Biomed. Inform., 77:34–49, Jan. 2018. doi: 10.1016/j.jbi.2017.11.011.

[30] J. Wei, Y. Tay, R. Bommasani, C. Raffel, B. Zoph, S. Borgeaud, D. Yogatama, M. Bosma, D. Zhou, D. Metzler, E. H. Chi, T. Hashimoto, O. Vinyals, P. Liang, J. Dean, and W. Fedus. Emergent abilities of large language models, 2022. URL https://arxiv.org/abs/2206.07682.

[31] T. Wolf, L. Debut, V. Sanh, J. Chaumond, C. Delangue, A. Moi, P. Cistac, T. Rault, R. Louf, M. Funtowicz, J. Davison, S. Shleifer, P. von Platen, C. Ma, Y. Jernite, J. Plu, C. Xu, T. Le Scao, S. Gugger, M. Drame, Q. Lhoest, and A. Rush. Transformers: State-of-the-art natural language processing. In Proceedings of the 2020 Conference on Empirical Methods in Natural Language Processing: System Demonstrations, pages 38–45, Stroudsburg, PA, USA, Oct. 2020. Association for Computational Linguistics. doi: 10.18653/v1/2020.emnlp-demos.6.

[32] Y. Xia, Y. Zhao, W. Wu, and S. Li. Debiasing Generative Named Entity Recognition by Calibrating Sequence Likelihood. In A. Rogers, J. Boyd-Graber, and N. Okazaki, editors, Proceedings of the 61st Annual Meeting of the Association for Computational Linguistics (Volume 2: Short Papers), pages 1137–1148, Toronto, Canada, July 2023. Association for Computational Linguistics. doi: 10.18653/v1/2023.acl-short.98. URL https://aclanthology.org/2023.acl-short.98/.

[33] F. Yan, P. Yu, and X. Chen. LTNER: Large language model tagging for named entity recognition with contextualized entity marking. In Antonacopoulos, A., Chaudhuri, S., Chellappa, R., Liu, CL., Bhattacharya, S., Pal, U., editor, Pattern Recognition,, Lecture Notes in Computer Science, page vol 15319. Springer, Apr. 2024. doi: 10.1007/978-3-031-78495-8_25.

[34] M. Yasunaga, J. Leskovec, and P. Liang. LinkBERT: Pretraining Language Models with Document Links. Proceedings of the 60th Annual Meeting of the Association for Computational Linguistics (Volume 1: Long Papers), Mar. 2022. doi: 10.18653/v1/2022.acl-long.551. URL http://arxiv.org/abs/2203.15827 2203.15827 [cs].

[35] U. Zaratiana, N. Tomeh, P. Holat, and T. Charnois. An Autoregressive Text-to-Graph Framework for Joint Entity and Relation Extraction. Proceedings of the AAAI Conference on Artificial Intelligence, 38(17):19477–19487, Mar. 2024. ISSN 2374-3468. doi: 10.1609/aaai.v38i17.29919. URL https://ojs.aaai.org/index.php/AAAI/article/view/29919.

[36] U. Zaratiana, N. Tomeh, P. Holat, and T. Charnois. GLiNER: Generalist model for named entity recognition using bidirectional transformer. In Proceedings of the 2024 Conference of the North American Chapter of the Association for Computational Linguistics: Human Language Technologies (Volume 1: Long Papers), pages 5364–5376, Stroudsburg, PA, USA, 2024. Association for Computational Linguistics. doi: 10.18653/v1/2024.naacl-long.300.

[37] J. Zhang, X. Liu, X. Lai, Y. Gao, S. Wang, Y. Hu, and Y. Lin. 2INER: Instructive and In-Context Learning on Few-Shot Named Entity Recognition. In H. Bouamor, J. Pino, and K. Bali, editors, Findings of the Association for Computational Linguistics : EMNLP 2023, pages 3940–3951, Singapore, Dec. 2023. Association for Computational Linguistics. doi: 10.18653/v1/2023.findings-emnlp.259. URL https://aclanthology.org/2023.findings-emnlp.259/.

[38] S. Zhou, N. Wang, L. Wang, H. Liu, and R. Zhang. CancerBERT: a cancer domain-specific language model for extracting breast cancer phenotypes from electronic health records. Journal of the American Medical Informatics Association : JAMIA, 29 (7):1208–1216, Mar. 2022. ISSN 1067-5027. doi: 10.1093/jamia/ocac040. URL https://www.ncbi.nlm.nih.gov/pmc/articles/PMC9196678/.

[39] W. Zhou, S. Zhang, Y. Gu, M. Chen, and H. Poon. UniversalNER: Targeted Distillation from Large Language Models for Open Named Entity Recognition. arXiv, 2024. doi: 10.48550/arXiv.2308.03279. URL https://arxiv.org/abs/2308.03279.

