## Supplementary File 1 for "Benchmarking LLM-based Information Extraction Tools for Medical Documents"

### Can AI-powered Information Extraction Tools Handle Medical Documents: Benchmark Study
### Supplementary Materials

Aaron Yu Jochen Weile Mélanie Courtot

October 20, 2026

#### Supplementary figures

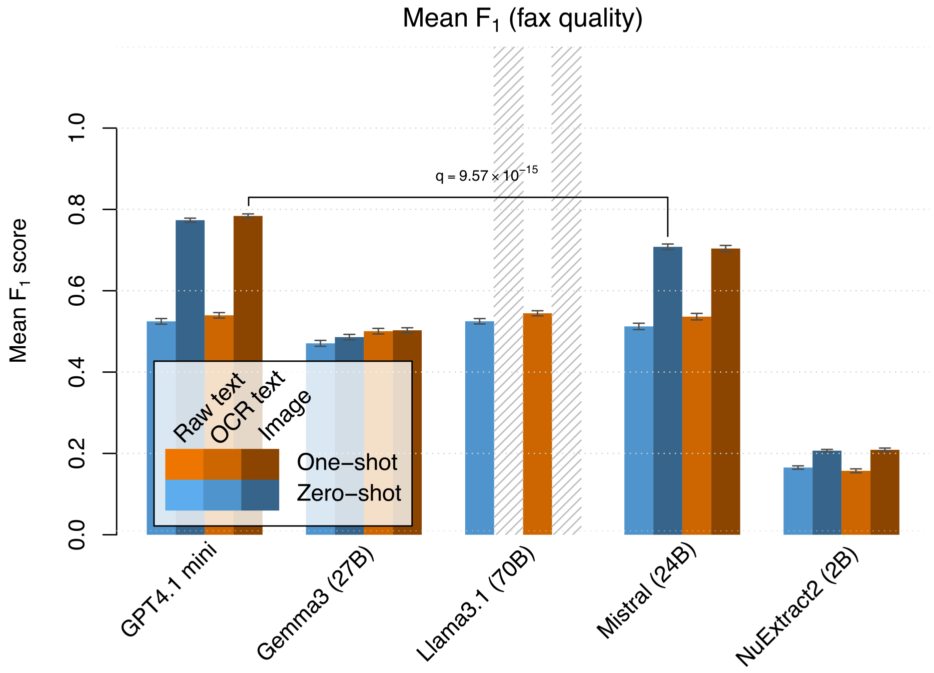


Figure S1: Mean $F_1$ scores across extractions from fax-distorted documents for each tool split by input modality and prompt type.

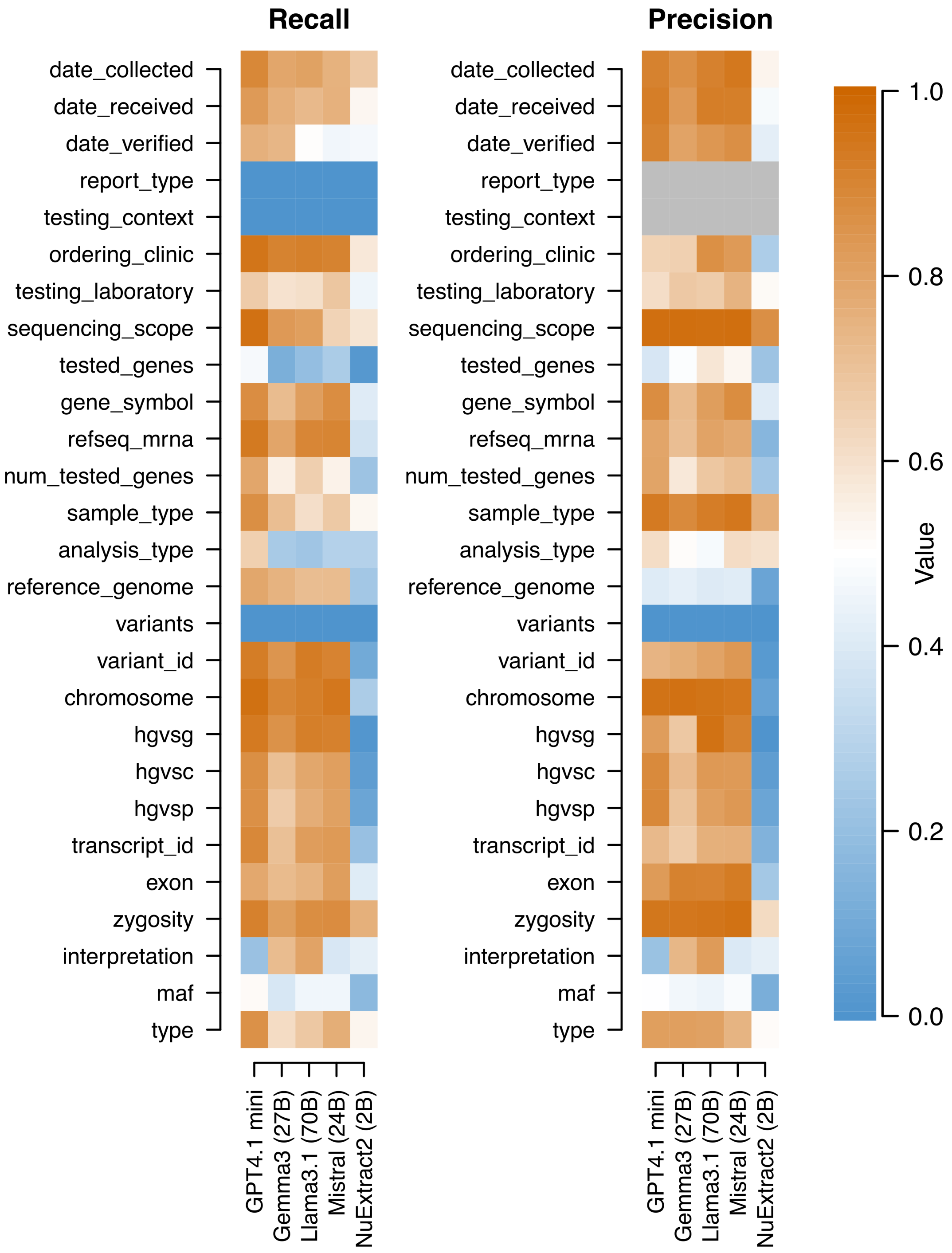


Figure S2: Recall and precision separated by prompted field for each tested tool across all parseable extraction outputs. Orange indicates high precision or recall, blue indicates low precision or recall. Gray indicates NaN due to division by zero, i.e. for recall: no relevant fields to be extracted in the document templates of parseable outputs; for precision: tool did not extract anything for that field. The `tested_genes` and `variants` fields are collections (dict of dict and list of dict, respectively). Their indicated recall and precision values represent the elements of those collections.

### Supplementary tables

Table S1: Literature review of existing information extraction (IE) tools based on large language models.

| Tool name | Publication | Runs out-of-box | Structured output | Sufficient context size | Open source | Biomedical applicability | Custom fields |
|---|---|---|---|---|---|---|---|
| NERRE | Dagdelen et al. [2024] | No | Yes | Yes | Yes | No (mat.science) | No |
| ATG | Zaratiana et al. [2024a] | No | Yes | Yes | No weights | Yes | No |
| BioGPT | Luo et al. [2022] | No | n/a | No | Yes | Yes | Yes |
| Bio-LinkBERT | Yasunaga et al. [2022] | No | Yes | Yes | Yes | Yes | Yes |
| GIX | Gill et al. [2024] | Yes | Yes | Yes | Yes | Protein interactions only | No |
| BERN2 | Sung et al. [2022] | Yes | Yes | Yes | Yes | Selected cell biology only | No |
| BioBERT | Lee et al. [2020] | No | Yes | Yes | Yes | Yes | Yes |
| GPT-NER | Wang et al. [2025] | No | Yes | Yes | Yes | Yes | No |
| Re-Rank-NER | Xia et al. [2023] | No | Yes | Yes | No | Yes | No |
| UniversalNER | Zhou et al. [2024] | No | Yes | Yes | Yes | Yes | No |
| CancerBERT | Zhou et al. [2022] | No | Yes | Yes | No | Yes | Yes |
| P-ICL | Jiang et al. [2024] | No | Yes | Yes | No | Yes | No |
| LTNER | Yan et al. [2024] | No | Yes | Yes | Yes | Yes | No |
| 2INER | Zhang et al. [2023] | No | Yes | Yes | No | Yes | No |
| NSSC | García-Barragán et al. [2025] | Yes | Yes | Yes | Yes | Spanish only | No |
| GliNER | Zaratiana et al. [2024b] | Yes | Yes | Yes | Yes | Yes | No |
| NuExtract 2.0 | Constantin et al. [2024] | Yes | Yes | Yes | Yes | Yes | Yes |

### Supplementary methods

#### The scoring system

The scoring system operates on the reference data (meant to be extracted), as well as the LLM or tool response containing the extractions. Both are provided as JSON dictionaries, with field names such as `date_collected`, `report_type` or `tested_genes`. Most of these fields contain simple string values, while a few can contain lists or dictionaries of sub-dictionaries. For example, the `variants` field typically contains a list of sub-dictionaries, which contain fields for each variant, such as `gene_symbol`, `transcript_id` or `zygosity`. However, not all of the document templates use all the possible fields, and some document templates even contain conflicting information, such as a hard-wired genetic test context. To account for these cases, we perform a masking step before scoring begins. Fields missing from a the given template are reset to an empty string in the reference dictionary, while conflicting fields are marked to be ignored during the scoring step. Furthermore, we distinguish between fields that require perfect matches (such as gene symbols, transcript IDs, variant IDs, etc. and fields that allow inexact matching, such as names of hospitals, testing laboratories, report types or testing contexts.

The scoring then proceeds as follows:

1. We identify any field names in the response that are not defined in the reference data. These are deemed to be hallucinations and tallied as false positives.

2. We then identify any field names in the reference data that are missing from the response. These are then tallied as false negatives.

3. Then, for each field name that matches between reference and response, we apply the following procedure:

(a) If the field has no reference value (due to being masked for being absent in the document template), then an empty response value counts as a true positive, while any other value for this field counts as a false positive.

(b) If the field has a simple string value in the reference data, then:

   i. If the response value is empty, it is tallied as a false negative.

   ii. Otherwise (when both reference and response have a simple string value), we normalize both strings, by converting both of them to upper-case characters and then applying a few rules that harmonize common variations (e.g. VUS = Variant of Uncertain Clinical Significance = Variant of Uncertain Significance).

   iii. If the field requires strict perfect matches (such as for gene names), then a perfect match is tallied as a true positive, while a mismatch is tallied as both a false negative (for failing to report the reference value) and a false positive (for providing a different incorrect value).

   iv. Otherwise, if inexact matching is allowed, then we perform a Needleman-Wunsch global character alignment of the two strings, count the number of matching, mismatching, inserted and deleted characters and then apply the following formulae to calculate partial true positives, false positives, and false negatives:

$$TP = \frac{\#\text{matches}}{\text{length(alignment)}}$$
$$FP = \frac{\#\text{mismatches} + \#\text{insertions}}{\text{length(extraction)}}$$
$$FN = \frac{\#\text{mismatches} + \#\text{deletions}}{\text{length(reference)}}$$

   . These equations are designed to harmonize with the exact matching approach above: A complete mismatch results in (i) no matching characters, (ii) all extracted characters being either insertions or mismatches, and (iii) all reference characters being either deletions or mismatches, thus counting as 0 TP, 1 FP and 1 FN.

(c) If the reference field does not have a simple string value, but instead contains a collection (such as a list of dictionaries (`variants`) or dictionary of dictionaries (`tested_genes`)), we must ensure that we compare the correct corresponding list entries to each other, as their sort order cannot be guaranteed, and the sizes of the two collections can differ. To solve this problem we apply the following procedure:

   i. We collect the IDs of all of the collection elements. In case of a dictionary we use the keys as IDs, whereas for lists, we construct an ID from suitable list entries (such as the gene name and hgvs variant descriptor).

   ii. We then calculate the pairwise dissimilarity between all possible combinations of element IDs. This is again done by performing a character alignment and counting the number of matches, mismatches, deletions and insertions. Then, the dissimilarity is calculated as $d = \#mismatches + \#insertions + \#deletions - \#matches$. Thus, a perfect match has a dissimilarity of $-(length(ID))$, whereas a complete mismatch has a dissimilarity of $\text{length(ID1)} + \text{length(ID2)}$.

   iii. We then greedily assign reference and response collection elements to each other starting with the least dissimilar pairing.

   iv. The values of the paired entries are themselves dictionaries containing information such as gene names or variant IDs. To score these dictionary entries, we recursively use the same procedure from steps 1-3.

### Prompt texts

#### Zero-shot prompt

You are a high-accuracy curator for clinical, genomic, and diagnostic data from germline lab report documents.
Your output must exactly follow a JSON format detailed below. Do not add extra fields, comments, explanations or summaries.
You are efficient and precise, extracting only the requested fields that can found in the provided document. If the document does not contain information for a requested field, do not guess and leave the field blank instead.

Please extract and return the following fields from the report document:

A. Clinical Testing Info
- Analysis Type - The type of analysis performed. For example: Variant analysis, Karyotyping, Microarray, etc.
- Sequencing Scope - The scope of the sequencing assay. Allowed values: "Gene panel", "Targeted variant testing", "Whole-Exome-Sequencing (WES)", "Whole-Genome-Sequencing (WGS)", "Whole-Transcriptome-Sequencing (WTS)"
- Tested Genes - The list of gene symbols that were tested in the assay
- RefSeq mRNA transcripts - The transcript identifier for the tested genes, in RefSeq format, e.g. "NM_000123.3"
- Sample Type - The type of sample used in the assay. For example: "Amplified DNA", "ctDNA", "Total DNA", "Total RNA"

B. Report Metadata
- Report Dates - The dates on which the samples were collected, received, verified, etc. Use the YYYY-MM-DD date format
- Report Type - The type of report. Allowed values: "Pathology" or "Molecular Genetics"
- Testing Context - The legal context in which the testing was performed: Allowed values: "Clinical" or "Research"
- Ordering Clinic - The name of the clinic who requisitioned the test. Include the city where the clinic is located when possible. (e.g. Mount Sinai Hospital (Toronto))
- Testing Laboratory - The name of the laboratory who performed the test. Include city if possible. (e.g. Ontario Institute for Cancer Research (Toronto))
- Reference genome - The reference genome assembly relative to which the variants are described (e.g. GRCh37)

C. Variant Details
- Variant ID - The OMIM, ClinVar, or dbSNP identifier for a variant. e.g. "VCV000055407.65"
- Gene Symbol - The HGNC symbol for the gene in which the variant is located. e.g. BRCA1
- Transcript ID - e.g., NM_000123.3
- HGVS descriptors - These descriptor strings encode the molecular basis of the variant in three contexts:
  - hgvsg: The genomic context (e.g. "g.43063903G>T"),
  - hgvsc: The coding sequence context: (e.g. "c.5123C>A"),
  - hgvsp: The context in the translated protein (e.g. "p.Ala1708Glu")
- Chromosome - the chromosome on which the variant is located, e.g. chr1, chr22, chrX, chrY
- Exon - The exon within the gene in which the variant is located
- Zygosity - Whether the variant was found in a homozygous, heterozygous, or compound heterozygous configuration.
- Interpretation - The ACMG clinical classification of the variant. Allowed values "Pathogenic", "Likely pathogentic", "Variant of uncertain significance (VUS)", "Likely benign", or "Benign"
- MAF - The minor allele frequency of the variant.
- Type - The molecular type of the variant, e.g. frameshift, nonsense, synonymous, missense

Your response must follow the below JSON format exactly:

‘‘‘json

```
{
  "date_collected": "",
  "date_received": "",
  "date_verified": "",
  "report_type": "",
  "testing_context": "",
  "ordering_clinic": "",
  "testing_laboratory": "",
  "sequencing_scope": "",
  "tested_genes": {
    "GENE1": {
      "gene_symbol": "GENE1",
```

````
      "refseq_mrna": ""
    }
  },
  "num_tested_genes": "",
  "sample_type": "",
  "analysis_type": "",
  "reference_genome": "",
  "variants": [
    {
      "gene_symbol": "",
      "variant_id": "",
      "chromosome": "",
      "hgvsg": "",
      "hgvsc": "",
      "hgvsp": "",
      "transcript_id": "",
      "exon": "",
      "zygosity": "",
      "interpretation": "",
      "maf": "",
      "type":""
    }
  ]
}
```

If the information for a given field could not be found in the document, leave the field blank (use "")
    . All fields must be included, even if empty. Do NOT add summaries or comments. Validate your
    output for format errors.
````

## One-shot prompt

````
You are a high-accuracy curator for clinical, genomic, and diagnostic data from scanned and potentially
     OCR-processed lab report documents.
Your output must exactly follow a specific JSON format detailed below. Do not add extra fields,
    comments, explanations or summaries.
Be efficient and precise, extracting only the requested fields that can found in the provided document.
     If the document does not contain information for a requested field, do not guess and leave its
    JSON value blank instead.

Please extract and return the following fields from the report document:

A. Clinical Testing Info
- Analysis Type - The type of analysis performed. For example: Variant analysis, Karyotyping,
    Microarray, etc.
- Sequencing Scope - The scope of the sequencing assay. Allowed values: "Gene panel", "Targeted variant
     testing", "Whole-Exome-Sequencing (WES)", "Whole-Genome-Sequencing (WGS)", "Whole-Transcriptome-
    Sequencing (WTS)"
- Tested Genes - The list of gene symbols that were tested in the assay
- RefSeq mRNA transcripts - The transcript identifier for the tested genes, in RefSeq format, e.g. "
    NM_000123.3"
- Sample Type - The type of sample used in the assay. For example: "Amplified DNA", "ctDNA", "Total DNA
    ", "Total RNA"

B. Report Metadata
- Report Dates - The dates on which the samples were collected, received, verified, etc. Use the YYYY-
    MM-DD date format
- Report Type - The type of report. Allowed values: "Pathology" or "Molecular Genetics"
- Testing Context - The legal context in which the testing was performed: Allowed values: "Clinical" or
     "Research"
- Ordering Clinic - The name of the clinic who requisitioned the test. Include the city where the
    clinic is located when possible. (e.g. Mount Sinai Hospital (Toronto))
- Testing Laboratory - The name of the laboratory who performed the test. Include city if possible. (e.
    g. Ontario Institute for Cancer Research (Toronto))
- Reference genome - The reference genome assembly relative to which the variants are described (e.g.
    hg38)

C. Variant Details
- Variant ID - The OMIM, ClinVar, or dbSNP identifier for a variant. e.g. "VCV000055407.65"
- Gene Symbol - The HGNC symbol for the gene in which the variant is located. e.g. BRCA1
- Transcript ID - e.g., NM_000123.3
- HGVS descriptors - These descriptor strings encode the molecular basis of the variant in three
    contexts:
````
````

~~~~
  - hgvsg: The genomic context (e.g. "g.43063903G>T"),
  - hgvsc: The coding sequence context: (e.g. "c.5123C>A"),
  - hgvsp: The context in the translated protein (e.g. "p.Ala1708Glu")
- Chromosome - the chromosome on which the variant is located, e.g. chr1, chr22, chrX, chrY
- Exon - The exon within the gene in which the variant is located
- Zygosity - Whether the variant was found in a homozygous, heterozygous, or compound heterozygous
    configuration.
- Interpretation - The ACMG clinical classification of the variant. Allowed values "Pathogenic", "
    Likely pathogentic", "Variant of uncertain significance (VUS)", "Likely benign", or "Benign"
- MAF - The minor allele frequency of the variant.
- Type - The molecular type of the variant, e.g. frameshift, nonsense, synonymous, missense

Your response must follow the below JSON format exactly:

```json
{
  "date_collected": "",
  "date_received": "",
  "date_verified": "",
  "report_type": "",
  "testing_context": "",
  "ordering_clinic": "",
  "testing_laboratory": "",
  "sequencing_scope": "",
  "tested_genes": {
    "GENE1": {
      "gene_symbol": "GENE1",
      "refseq_mrna": ""
    }
  },
  "num_tested_genes": "",
  "sample_type": "",
  "analysis_type": "",
  "reference_genome": "",
  "variants": [
    {
      "gene_symbol": "",
      "variant_id": "",
      "chromosome": "",
      "hgvsg": "",
      "hgvsc": "",
      "hgvsp": "",
      "transcript_id": "",
      "exon": "",
      "zygosity": "",
      "interpretation": "",
      "maf": "",
      "type":""
    }
  ]
}
```

If the information for a given field could not be found in the document, leave the field blank (use "")
    . All fields must be included, even if empty. Do NOT add summaries or comments. Validate your
    output for format errors.

Here is an example of a document and its correct extraction:

<example>
<document>
Draft--- Do not use for patient care decisions
------------------------------------------------------------------------

  ---------- ------------------------------------------------------------ --
  **CHEO**   **Genetics Diagnostic Laboratory**\
             401 Smyth Rd, Ottawa, Ontario K1H 8L1\
             (Phone) 613-737-7600 ext. 3796 $\cdot$ (Fax) 613-738-4814

  ---------- ------------------------------------------------------------ --

  --------------- ---------- --------------- ----------
  **Patient:**    Redacted   **Location:**   Redacted
  **DOB/Sex:**    Redacted   **Pedigree:**   Redacted
~~~~

| | |
|---|---|
| **CHEO MRN:** | Redacted |

Hereditary Breast/Ovarian/Prostate Cancer Panel (In process)

---

| | | | |
|---|---|---|---|
| Authorized by: | Redacted | Collected: | 2021-08-17 07:24:43 |
| Ordered by: | Sinai Health System (Toronto) | Received: | 2021-08-18 14:50:44 |
| ID: | Redacted | Verified On: | 2021-08-19 10:28:31 |

### Specimens

**A** - Hereditary Breast/Ovarian/Prostate Cancer Panel, Genetics Blood, edta x2

### Reason for Referral

(GD1) \> or = 5% likelihood of Pathogenic/Likely Pathogenic variant in affected or unaffected individual

### MOLECULAR GENETIC RESULT

Summary of Results: **A pathogenic variant detected.**

Finding(s):

| **Gene** | **Variant (c.)** | **Amino acid change (p.)** | **Zygosity** | **Interpretation** |
|---|---|---|---|---|
| ARID5B | c.2569C\>A | p.Pro857Thr | heterozygous | Pathogenic |

### INTERPRETATION

A pathogenic variant was detected in this individual. The interpretation of this result is summarized below.

The c.2569C\>A variant in ARID5B It was identified in 1/250010 (0.0004%) of alleles tested from control populations in the Genome Aggregation Database (gnomAD). To the best of our knowledge, it has not been previously reported in the literature. The Pro857 residue is weakly conserved in evolution. In silico analysis programs (SIFT, PolyPhen-2, Mutation Taster) predict this variant not to be tolerated . This variant is listed in ClinVar (VCV007921369). In accordance with existing evidence, this variant is therefore classified as a pathogenic variant.

Please refer to the ClinVar database for further infomation about this variant(s). If a formal variant re-evaluation is required, please contact our laboratory.

No other clinically relevant sequence variants or large deletions or duplications were detected in the genes tested (see the Test Details section). Although no other pathogenic variants have been found, as the analytical sensitivity of this analysis is based on results of validation studies performed by our laboratory is $\sim 99\%$, it is possible that a pathogenic variant not detectable by these assays is present in one of these genes. In addition; as this test does not detect all clinically significant variants associated with this disorder, it is also possible that a pathogenic variant in another gene not tested is present in this individual.

Genes included in this panel are associated with an increased risk for certain types of hereditary cancers. While the majority of cancers occur sporadically, a proportion may be due to inherited pathogenic variants in genes associated with cancer. Pathogenic variants in these genes are associated with an increased susceptibility to certain malignancies, autosomal recessive conditions, and may cause other non-cancer related symptoms. It is important to note that not all individuals with a pathogenic variant will develop cancer. Genetic testing of at-risk first degree relatives should be considered once identification of a variant has occurred in a proband. The reproductive implications of being a carrier of a pathodenic variant should be discussed with an appropriate health care professional. Genetic counselling is recommended concerning the implications of these results to this individual and this individual's family.

This assay is based on the current state of knowledge of the genetic basis of this disorder and designed to identify constitutional genetic changes in the tested gene/loci (see Test Details). The possibility of null alleles due to rare family specific variants in the primer/probe binding sites cannot be completely ruled out. Family relationships are assumed as stated and no attempt has been made to verify these relationships. Laboratory results are subject to approximately 0.5% error in any of the pre-analytical, analytical or post-analytical phases of the test \[Clin. Chem 48(5).691-698\].

### Methodology Used

Next Generation Sequencing HEREDITARY BREAST AND OVARIAN CANCER 5 gene panel, HOXB13 c.251G\>A (p.Gly84Glu) variant sequencing, and MLPA for a subset of genes

Electronically signed by \[Not yet signed\] on \[Date\] at \[Time\]

### Test Details

Genes analyzed: MDC1, CUX1, ARID5B, TCEA1, POTEF,

Genomic DNA is enriched for targeted regions using custom designed capture probes (KAPA HyperChoice) and a hybridization-based protocol (KAPA HyperPlus Custom Library, KAPA Biosystems-Roche) followed by next generation sequencing (NGS) on a NextSeq platform (Illumina). Read alignment and single nucleotide variants (SNV) and small insertion/deletions (indels) calling are performed using NextGENe software (SNP/INDEL analysis, SoftGenetics) SNVs and indels within coding exons and 20 bps flanking regions are analyzed. Certain known likely pathogenic or pathogenic variants outside these reqions are also included in the analysis (for a complete list please contact our laboratory). Exonic deletions and duplications in all targeted genes (except PMS2) are called using an in-house copy number variant (CNV) analysis algorithm (PMID 27376475). Multiplex ligation-dependent probe amplification (MLPA) is performed for CNV analysis for PMS2. Note that some MLPA kits used as part of this test may contain probes for genes or variants that are not included in our test panels, which could result in incidental findings. All reported variants are confirmed by a second technology including Sanger sequencing; MLPA, long range PCR, or qPCR. Based on validation study results, this assay achieves 99% analytical sensitivity and specificity for SNV, indels \<10bp in length and exonic deletions and duplications; however, sensitivity for indels larger than 10bp but smaller than a full exon may be reduced. This assay is not designed and validated for detection of mosaicism. The following genes and corresponding exons are prone to misalignment due to presence of highly homologous regions in the genome and therefore, are at increased risk of false results: ATM exon 28, CHEK2 exons 11-15, PTEN exon 9 and PMS2 exons 1-5, 9, 11-15 (PMID 27228465). Variant nomenclature is based on the Human Genome Variation Society guidelines. Variant interpretation and classification is based on published guidelines (PMID 25741868). Only pathogenic, likely pathogenic, and variants of uncertain clinical significance are reported.

The following transcripts are used in this analysis: MDC1 (NM_014641.3), CUX1 (NM_181552.4), ARID5B (NM_032199.3), TCEA1 (NM_006756.4), POTEF (NM_001099771.2),

*CNV analysis only; \^single variant c.251G\>A, p.(Gly84Glu) only

</document>

<extraction>

{
    "date_collected": "2021-08-17 07:24:43",
    "date_received": "2021-08-18 14:50:44",
    "date_verified": "2021-08-19 10:28:31",
    "report_type": "Molecular Genetics",
    "testing_context": "",
    "ordering_clinic": "Sinai Health System (Toronto)",

```
    "testing_laboratory": "CHEO (Ottawa)",
    "sequencing_scope": "Gene panel",
    "tested_genes": {
        "MDC1": {
            "gene_symbol": "MDC1",
            "refseq_mrna": "NM_014641.3"
        },
        "CUX1": {
            "gene_symbol": "CUX1",
            "refseq_mrna": "NM_181552.4"
        },
        "ARID5B": {
            "gene_symbol": "ARID5B",
            "refseq_mrna": "NM_032199.3"
        },
        "TCEA1": {
            "gene_symbol": "TCEA1",
            "refseq_mrna": "NM_006756.4"
        },
        "POTEF": {
            "gene_symbol": "POTEF",
            "refseq_mrna": "NM_001099771.2"
        }
    },
    "num_tested_genes": 5,
    "sample_type": "Blood",
    "analysis_type": "Genetics",
    "variants": [
        {
            "gene_symbol": "ARID5B",
            "variant_id": "VCV007921369",
            "chromosome": "",
            "hgvsg": "",
            "hgvsc": "c.2569C>A",
            "hgvsp": "p.Pro857Thr",
            "type": "missense",
            "transcript_id": "",
            "exon": "",
            "zygosity": "heterozygous",
            "interpretation": "Pathogenic",
            "maf": "0.0004%"
        }
    ],
    "reference_genome": ""
}

</extraction>
</example>
```


### References


A. Constantin, L. Cripwell, N. Fradet, S. Dréano, and E. Bernard. Nuextract: A foundation model for structured extraction, June 2024. URL https://github.com/numindai/nuextract. original-date: 2025-02-18T14:07:59Z.

J. Dagdelen, A. Dunn, S. Lee, N. Walker, A. S. Rosen, G. Ceder, K. A. Persson, and A. Jain. Structured information extraction from scientific text with large language models. *Nature Communications*, 15 (1):1–14, Feb. 2024. ISSN 2041-1723. doi: 10.1038/s41467-024-45563-x. URL https://www.nature.com/articles/s41467-024-45563-x.

Á. García-Barragán, A. Sakor, M.-E. Vidal, E. Menasalvas, J. C. S. Gonzalez, M. Provencio, and V. Robles. NSSC: a neuro-symbolic AI system for enhancing accuracy of named entity recognition and linking from oncologic clinical notes. *Medical & Biological Engineering & Computing*, 63(3):749–772, 2025. ISSN 0140-0118. doi: 10.1007/s11517-024-03227-4. URL https://www.ncbi.nlm.nih.gov/pmc/articles/PMC11891111/.

J. K. Gill, M. Chetty, S. Lim, and J. Hallinan. Large language model based framework for automated extraction of genetic interactions from unstructured data. *PLOS ONE*, 19(5):e0303231, May 2024. ISSN 1932-6203. doi: 10.1371/journal.pone.0303231. URL `https://journals.plos.org/plosone/article?id=10.1371/journal.pone.0303231`.

G. Jiang, Z. Ding, Y. Shi, and D. Yang. P-ICL: Point in-context learning for named entity recognition with large language models. *ArXiv*, abs/2405.04960, May 2024. doi: 10.48550/arXiv.2405.04960.

J. Lee, W. Yoon, S. Kim, D. Kim, S. Kim, C. H. So, and J. Kang. BioBERT: a pre-trained biomedical language representation model for biomedical text mining. *Bioinformatics*, 36(4):1234–1240, Feb. 2020. ISSN 1367-4803. doi: 10.1093/bioinformatics/btz682. URL `https://doi.org/10.1093/bioinformatics/btz682`.

R. Luo, L. Sun, Y. Xia, T. Qin, S. Zhang, H. Poon, and T.-Y. Liu. BioGPT: generative pre-trained transformer for biomedical text generation and mining. *Briefings in Bioinformatics*, 23(6):bbac409, Nov. 2022. ISSN 1477-4054. doi: 10.1093/bib/bbac409. URL `https://doi.org/10.1093/bib/bbac409`.

M. Sung, M. Jeong, Y. Choi, D. Kim, J. Lee, and J. Kang. BERN2: an advanced neural biomedical named entity recognition and normalization tool. *Bioinformatics*, 38(20):4837–4839, Sept. 2022. ISSN 1367-4803. doi: 10.1093/bioinformatics/btac598. URL `https://www.ncbi.nlm.nih.gov/pmc/articles/PMC9563680/`.

S. Wang, X. Sun, X. Li, R. Ouyang, F. Wu, T. Zhang, J. Li, G. Wang, and C. Guo. GPT-NER: Named entity recognition via large language models. In *Findings of the Association for Computational Linguistics: NAACL 2025*, pages 4257–4275, Stroudsburg, PA, USA, 2025. Association for Computational Linguistics. doi: 10.18653/v1/2025.findings-naacl.239.

Y. Xia, Y. Zhao, W. Wu, and S. Li. Debiasing Generative Named Entity Recognition by Calibrating Sequence Likelihood. In A. Rogers, J. Boyd-Graber, and N. Okazaki, editors, *Proceedings of the 61st Annual Meeting of the Association for Computational Linguistics (Volume 2: Short Papers)*, pages 1137–1148, Toronto, Canada, July 2023. Association for Computational Linguistics. doi: 10.18653/v1/2023.acl-short.98. URL `https://aclanthology.org/2023.acl-short.98/`.

F. Yan, P. Yu, and X. Chen. LTNER: Large language model tagging for named entity recognition with contextualized entity marking. In Antonacopoulos, A., Chaudhuri, S., Chellappa, R., Liu, CL., Bhattacharya, S., Pal, U., editor, *Pattern Recognition,*, Lecture Notes in Computer Science, page vol 15319. Springer, Apr. 2024. doi: 10.1007/978-3-031-78495-8_25.

M. Yasunaga, J. Leskovec, and P. Liang. LinkBERT: Pretraining Language Models with Document Links. *Proceedings of the 60th Annual Meeting of the Association for Computational Linguistics (Volume 1: Long Papers)*, Mar. 2022. doi: 10.18653/v1/2022.acl-long.551. URL `http://arxiv.org/abs/2203.15827`. arXiv:2203.15827 [cs].

U. Zaratiana, N. Tomeh, P. Holat, and T. Charnois. An Autoregressive Text-to-Graph Framework for Joint Entity and Relation Extraction. *Proceedings of the AAAI Conference on Artificial Intelligence*, 38(17):19477–19487, Mar. 2024a. ISSN 2374-3468. doi: 10.1609/aaai.v38i17.29919. URL `https://ojs.aaai.org/index.php/AAAI/article/view/29919`.

U. Zaratiana, N. Tomeh, P. Holat, and T. Charnois. GLiNER: Generalist model for named entity recognition using bidirectional transformer. In *Proceedings of the 2024 Conference of the North American Chapter of the Association for Computational Linguistics: Human Language Technologies (Volume*

*1: Long Papers)*, pages 5364–5376, Stroudsburg, PA, USA, 2024b. Association for Computational Linguistics. doi: 10.18653/v1/2024.naacl-long.300.

J. Zhang, X. Liu, X. Lai, Y. Gao, S. Wang, Y. Hu, and Y. Lin. 2INER: Instructive and In-Context Learning on Few-Shot Named Entity Recognition. In H. Bouamor, J. Pino, and K. Bali, editors, *Findings of the Association for Computational Linguistics : EMNLP 2023*, pages 3940–3951, Singapore, Dec. 2023. Association for Computational Linguistics. doi: 10.18653/v1/2023.findings-emnlp.259. URL https://aclanthology.org/2023.findings-emnlp.259/.

S. Zhou, N. Wang, L. Wang, H. Liu, and R. Zhang. CancerBERT: a cancer domain-specific language model for extracting breast cancer phenotypes from electronic health records. *Journal of the American Medical Informatics Association : JAMIA*, 29(7):1208–1216, Mar. 2022. ISSN 1067-5027. doi: 10.1093/jamia/ocac040. URL https://www.ncbi.nlm.nih.gov/pmc/articles/PMC9196678/.

W. Zhou, S. Zhang, Y. Gu, M. Chen, and H. Poon. UniversalNER: Targeted Distillation from Large Language Models for Open Named Entity Recognition. *arXiv*, 2024. doi: 10.48550/arXiv.2308.03279. URL https://arxiv.org/abs/2308.03279.
