## Supplementary File 2 for "Benchmarking LLM-based Information Extraction Tools for Medical Documents"

### DMSC

#### LABORATORY MEDICINE DIVISION

Medical Genetics Lab
123 Sample St, Doeford, AB, A1B 2C3

| | | | | | |
|---|---|---|---|---|---|
| **Patient Name:** Redacted | | | **DoB:** Redacted | | **Sex:** Redacted |
| **HC#:** Redacted | | **Location:** Redacted | **Alt: Hosp#:** Redacted | | **CR#:** Redacted |

| | | | |
|---|---|---|---|
| **Accession #:** | Redacted | **Referring Physician:** | Redacted |
| **Collection Date/Time** | 2021-07-19 02:48:08 | | |
| **Arrival Date/Time:** | 2021-07-20 12:43:12 | | |
| **Report Print Date:** | 2021-07-23 00:53:17 | cc: | |

Reason for Referral: HEREDITARY CANCER

Sample Type: Blood

Mock Specimen

#### RESULT

TEST DONE: Hereditary Breast/Ovarian Prostate Cancer Panel, 14 genes (FAM46C, NCKIPSD, HOOK3, PRRX1, ETV4, FLI1, ZEB2, DDB2, GRM3, MBD1, CD79B, CUL4B, DDX3X, FEV, )

RESULTS: A variant was detected in the FAM46C gene.

NM_017709.3(FAM46C):[c.38C>G(p.Ser13Cys)]:[=]

INTERPRETATION:
This missense variant in the FAM46C gene results in a single base substitution at position 38 of the coding sequence, which replaces a serine at position 13 with a cysteine . This modification may alter protein function. The variant has been reported multiple times in ClinVar, with conflicting interpretations. The variant has been observed in gnomAD with a frequency of 0.00193049 . This variant is classified as Variant of uncertain clinical significance .

Interpretation of these findings must be made in light of the clinical history and evaluation of this individual. Genetic counselling and appropriate clinical follow up are recommended for this individual.

DISEASE INFORMATION:
Hereditary cancer syndromes are associated with predisposition for a variety of cancers. The gene panel used in this analysis interrogates the coding regions of the genes specified above that have been associated with an increased risk for this syndrome. The finding of pathogenic or likely pathogenic variants in these genes can impact clinical management of individuals, and may support mutation-specific testing for at risk family members.

METHOD:
Testing was performed by massively parallel sequencing using a custom Roche KAPA HyperCap panel with target enrichment probes specific to 75 genes (listed below) with filtered analysis specific to the genes listed above. A library was generated from genomic DNA that included all coding regions of the panel genes indicated, including 20 bases past each intron/exon boundary, with the exception of certain genes where either copy number only (EPCAM, GREM1) or hotspot examination only (EGFR, HOXB13 , MITF) was done: The library was sequenced to an average minimum depth of 500 fold per amplicon, with a minimum required depth of 20 fold per nucleotide. Massively parallel sequencing has an analytic sensitivity and specificity of ~98% for the detection of nucleotide base alterations and small deletions and insertions. Large deletions or insertions were detected by comparative analysis of library sequencing read depths. The analytic sensitivity for the detection of these larger changes is estimated at 95%, and specificity at ~98%.

Pathogenic, likely pathogenic and variants of uncertain significance are reported when they occur in coding regions, variants of unknown significance are not reported when they occur in non-coding regions. Benign and likely benign variants are not reported, but can be provided upon request.

As required, pathogenic, likely pathogenic, and unclassified variants may have been confirmed by bidirectional Sanger sequencing or quantitative PCR with an analytic sensitivity and specificity of >99% for confirmation analyses. Copy number variants affecting the following genes may be masked due to the presence of multiple pseudogenes: ATM, BMPR1A, CHEK2, FH, NFI, PMS2, PTEN, RPS20, SDHC, SDHD, SMARCE1 .

If analysis of these regions is specifically required, please inform the lab so that additional studies can be discussed.

The nomenclature used is based on the Human Genetic Variation Society (HGVS) guidelines using the GRCh37/hg19 build and NCBI reference sequence database accession numbers as follows: AIP (NM_003977.3); APC (NM 001127510.2); ATM (NM_000051.3); AXIN2 (NM_004655.3); BAP1 (NM_004656.3); BARD1 (NM_000465.4); BMPR1A (NM_004329.2); BRCA1 (NM_007294.3); BRCA2 (NM_00005933); BRIP1 (NM_032043.3); CDC73 (NM_024529.4); CDHI (NM 004360.4); CDKA (NM_000075.3); CDKN1B (NM_004064.4); CDKNZA (NM_000077.4); CHEKZ (NM_007194.4); CTNNA1 (NM_001903.5); DICER1 (NM_177438.2); EGFR (NM_005228.3); EGLN1 (NM_022051.2); EPCAM (NM_002354.2)-CNV analysis only; EXT1 (NM_000127.2); EXT2 (NM_001178083.1); FLCN (NM_144997.7); FH (NM_000143.3); GALNT12 (NM_024642.5); GREM1 (NM_013372.6)-CNV analysis only; HOXB13 (NM_006361.5); KIT (NM_000222.2); LZTR[illegible] ([illegible]M_006767.4); MAX (NM 002382.4); MEN1 (NM_000244.3); MET (NM 001127500.2); MITF (NM_198159.2); MLH[illegible] (NM_[illegible]0249.3); MSH2 (NM_000251.2); MSHH (NM_002439.5); MSH6 (NM_000179.2); MUTYH (NM 001128425[illegible] NB[illegible] (NM_002485.4); NF1 (NM_001042492.2): NF2 (NM_000268.3); NTHL1 (NM_002528.7); PALB2 (NM_024675.3); [illegible]DG[illegible]RA ([illegible]NM_006206.5); PMS2 (NM_000535.6); POLD1 (NM_001256849.1); POLE (NM_006231.3); POT1 (NM_015450.[illegible] PR[illegible]AR1A (NM_212471.2); PTCH1 (NM_000264.4); PTEN (NM_000314.8); RAD51C (NM_058216.3); RAD51D ([illegible]8.3); RBI (NM_000321.2); RECQL (NM_002907.4); RET (NM_020975.5); RNF43 (NM_017763.5); RPS20 [illegible]2.3); SDHA (NM_004168.4); SDHAF2 (NM_017841.2); SDHB (NM_003000.3); SDHC (NM_003001.3)[illegible] SDH[illegible] (NM_[illegible]3002.4); SMAD4 (NM_005359.5); SMARCA4 (NM_ 001128849.1); SMARCB1 (NM_003073.4); SMARCE1 ([illegible].4); STK11 (NM_000455.4); SUFU (NM_016169.3); TMEM127 (NM_017849.3); TP53 (NM_000546.5); TS[illegible] ([illegible]368.5); TSC2 (NM_000548.3); VHL (NM_000551.3)

DISCLAIMER:
Library construction, sequencing, data analysis and confirmato[illegible]testin[illegible] was performed within the Medical Genetics Lab at Doeford Medical Science Campus, which is accredited [illegible] to perform such high complexity testing.

Reported b[illegible]
Redact[illegible]
Directo[illegible]al [illegible]etics Lab

[illegible]lectronically signed by:
[illegible]edacted
D[illegible]e Authorized: -

Mock spec[illegible]

CCH
Chimwald Children's Hospital
123 Mock St.
Chiemwald, ON A1B 2C3

**Molecular Diagnostics Lab**
**Department of Paediatric Laboratory Medicine**
CLIA ID: REDACTED
www.cch.ca/molgen/

#### COMPREHENSIVE CANCER TEST REPORT

| **Primary Tumour Type:** | **Tumour Cellularity:** | **Reason for Referral:** |
|---|---|---|
| Hemangiosarcoma | - | Tumour and Germline testing |

**Family History**: No known family history of cancer

**Previous Testing**: -

**Other case details**: -

##### SUMMARY OF SOMATIC AND GERMLINE FINDINGS:

**Somatic Findings:**

| | | |
|---|---|---|
| Tumour Mutation Burden: | Not actionable | 0.838 mutations/Mb |
| **SNV/indels and copy number changes** | Gene Name | Variant |
| Clinically actionable variants in patient's tumour type | - | - |
| Clinically actionable variants in a different tumour type | - | - |
| Clinically actionable variants detected at low VAF | - | - |
| Oncogenic variants with uncertain clinical actionability | EGFR | c.570T>C (p.Cys190=) |
| Variants of uncertain clinical significance | - | - |

**Germline Findings:**

| **SNV/indels and copy number changes** | Gene Name | Variant |
|---|---|---|
| Pathogenic/Likely pathogenic variants | - | - |
| Variants of uncertain significance | - | - |

Tumour DNA: An oncogenic variant with uncertain clinical actionability in the EGFR gene was identified.Tumour mutation burden is not actionable.

Germline DNA: The results suggest that these variants are mosaic (~39%) in the peripheral blood of this patient.

Mock Specimen

Note: For clinical use only. Results not generated in a forensically accredited laboratory

CCH
Chimwald Children's Hospital
123 Mock St.
Chiemwald, ON A1B 2C3

**Molecular Diagnostics Lab**
**Department of Paediatric Laboratory Medicine**
CLIA ID: REDACTED
www.cch.ca/molgen/

#### RESULTS:

**SOMATIC SEQUENCE Results:** Variants listed below were acquired in the patient tumour and detected at a VAF **at or above** the sensitivity for clinical testing:

- Fresh frozen samples >5% for SNVs; >10% for indels
- FFPE samples >10% for SNVs and Indels

**Tumour Mutation Burden:** Not actionable 0.838 mutations/Mb

**Tumour Mutation Burden (TMB):**
TMB was determined to be 0.838 mutations/Mb and therefore is not clinically actionable.

Tumours with high TMB may benefit from checkpoint inhibitor immunotherapy and are considered clinically actionable if they meet the eligibility criteria for open clinical trials (PMID: 27001570, 29658845). Refer to the Test Information section at the end of this report for details on TMB calculation.

| Gene Name/Transcript ID | Sequence variant (Effect on Protein) | Genomic Location | Variant Allele Fraction (%) | Clinical Actionability* |
|---|---|---|---|---|
| EGFR | c.570T>C | Chr7(GRCh38): | 75.54 | Variant of uncertain clinical significance |
| NM_005228.5 | (p.Cys190=) | g.55019586T>C | | |

*See 'Test Information' section of the report

#### SOMATIC VARIANT INTERPRETATION:

**Clinically actionable:** Variants with known therapeutic; prognostic or diagnostic actionability in the patients tumour type or in a different tumour type

No variants found.

**Variants of Uncertain Clinical Significance (VUS):** Variants of uncertain association with therapeutic, prognostic or diagnostic actionability.

Variant: c.570T>C (p.Cys190=) in the EGFR gene
Variant type:
Variant Allele Fraction: 75.54%
The c.570T>C variant located in exon 24of the EGFR gene has been reported twice in other tumors (Cosmic 1x, cBioPortal 1x). Loss of EGFR expression is associated with increased tumor growth, including in angiosarcomas ( PMIDs: 17893028, 14963148, 10365667). However; clinical actionability of this variant remains uncertain at this time.

**SOMATIC COPY NUMBER Results**: Copy number gain/loss variants listed below were acquired in the patients tumour

No variants to report

**GERMLINE SEQUENCE Results:** Variants listed below were found in the germline of this patient

Mock Specimen

CCH
Chimwald Children's Hospital
123 Mock St.
Chiemwald, ON A1B 2C3

Molecular Diagnostics Lab
Department of Paediatric Laboratory Medicine
CLIA ID: REDACTED
www.cch.ca/molgen/

| Gene Name Transcript ID | Sequence variant (Effect on Protein) | Genomic Location | Population Frequency (gnomAD all MAF%) | Zygosity | Inheritance pattern | Interpretation |
|---|---|---|---|---|---|---|
| No variants to report. | | | | | | |

#### GERMLINE VARIANT INTERPRETATION:

**GERMLINE COPY NUMBER Results:** Copy number gain/loss variants listed below were found in the germline of this patient.

No variants to report

**TEST INFORMATION**

**METHODS:**

The Comprehensive Cancer Panel uses Next-Generation sequencing (NGS) to analyze 13 genes associated w[illegible]h a w[illegible] spectrum of pediatric cancer and/or an increased risk for cancer predisposition: The complete list of genes o the panel is provided at the end of this r[illegible] Appen[illegible]. Genetic variants (SNVs and indels) from the coding sequences and intron/exon boundaries (±10 base pairs of intronic sequence), a[illegible] well [illegible] copy n[illegible]ber (CN) variants, were investigated for the Cancer Panel.

This test is performed by Next-Generation sequencing using Agilent SureSelect capture followed by paire[illegible]d se[illegible]ncing of the coding and splice site regions using the Illumina sequencing platform. Germline variant calls are generated using Genomic Analy[illegible] Kit [illegible]ATK) after read alignment with the Burrows-Wheeler Aligner (BWA) to genome build CRCh37/UCSC hg19 with decoy. Somatic substitution and in[illegible] calling w[illegible]re [illegible]rformed on matched tumour-normal samples using Mutect and MuTect2, respectively. CN calling was performed using the NxClinical software (v6.[illegible] BioDisc[illegible]ry)- Custom filtering was developed to prioritize variants.

Tumour Mutation Burden (TMB) is measured as the number of somatic variants (onl[illegible] SNVs [illegible]ithin [illegible]nic coding regions per Megabase (Mb) of DNA (VAF >10%, within ±10 base pairs of intron-exon junctions): TMB is considered 'high' when me[illegible]ur[illegible] to b[illegible] at least 10 mutationsIMb ad can be associated with exposure to mutagens such as UV light, microsatellite instability or abnormal activity in DNA dama[illegible]d rep[illegible]r pathways such as gemline or somatic variants in mismatch repair genes or in the proofreading domains of the POLE and POLDI replication [illegible] 28420421, 29056344) Tumours with high TMB may benefit from checkpoint inhibitor immunotherapy and are considered clinically actiona[illegible] if they [illegible]eet the criteria for open clinical trials (PMID: 27001570, 29658845). These criteria may evolve depending on the tumour type and expected median [illegible] reporte[illegible] in the scientific literature.

CN changes that are focal gains/losses (25 Mb) in clinically signific[illegible] genes c[illegible]y this panel will be reported. CN state is defined as: gain (23 copies), loss (~1 copy), amplification (25-10 copies, dependent on clinical actionab[illegible]y of [illegible]ne) [illegible]d homozygousibi-allelic loss (<1 copy). Sufficient genome-wide coverage allows for detection of larger segmental (>5 Mb), chromosome ar level a[illegible] c[illegible]omos[illegible]e gains/losses, as well as regions with copy neutral LOH (>10 Mb) However, exact breakpoints will require additional confirmation when located within o[illegible] target regions.

This analysis is based on current knowledge of [illegible] mole[illegible]lar genetics of the tumour type indicated on the requisition; Test results should only be used in conjunction with the patients clinical history, personal/family h[illegible]ry [illegible] cancer and any previous analysis of appropriate family members Unless specifically stated, it is assumed that family relationships are 3s indicated and that the [illegible] of individuals is correct. It is recommended that these test results be communicated t0 the patient in a setting that includes appropriate counsell[illegible]

**LIMITATIONS AND OTHER TEST NOTES**

**Abbreviations:**

AD - Autosomal Domin[illegible]
AR - Autosomal Recess[illegible]
CN - Copy Number
FF - Fresh Fr[illegible]
FFPE - Forma[illegible] [illegible]raffi[illegible] Embe[illegible]ded
HET - Heterozyg[illegible]
HEMI - Hemizygous
HOM - Homozygous
LOH - Loss of Heterozyg[illegible]ity
MAF - Minor Allele Frequency
TMB - Tumour Mutation Burden
VAF - Variant Allele Fraction
VUS - Variant of Uncertain Significance
UTR - Untranslated region
XL - X-linked

Clinical actionability refers to the clinical utility and validity of the variant in the context of diagnosis, prognosis or therapeutic response (sensitivity or resistance).

Germline variants are classified using the criteria outined by the American College of Medical Genetics (ACMG) guidelines (Richards et al: (2015) PMID: 25741868). Somatic variants are classified using the criteria outlined below:

**Therapeutic:** variant is therapeutically actionable

- Class 1: FDA-approved therapies show response in this (A) or another (B) tumour type
- Class 2: well-powered clinical trials show response or resistance in this (A) or another (B) tumour type

Molecular Diagnostics Lab
Department of Paediatric Laboratory Medicine
CLIA ID: REDACTED
www.cch.ca/molgen/

- Class 3: a small series, case reports show response or resistance in this (A) or another (B) tumour type; or the variant meets the inclusion criteria for a clinical trial in this (A) or another (B) tumour type

**Diagnostic**: variant is diagnostic of the patient's tumour type **Prognostic**: variant is prognostic in the patient's tumour type **Variants of Uncertain Significance (VUS)**: clinical actionability of this variant is currently uncertain; however, the variant may be biologically relevant in the patient's tumour type. There are four categories of VUSs:

- V1: well-characterized oncogenic variant (gain-of-function or loss-of-function) based on experimental or clinical evidence
- V2: Inferred oncogenic variant (loss-of-function) based on mutation type (i.e. stop gain or frameshift) or recurrence in cancer samples (e.g hotspot mutations that have not been formally characterized)
- V3: potentially oncogenic variant based on the gene that it is affecting and/or predicted impact on protein function (e.g- mutation [illegible] functional domain, causes aberrant gene expression, protein loss, pathway dysregulation or results in a mutational signature or hypermutation)
- VUS: there is currently insufficient published data to determine the significance of this variant, but it is in a gene where there is [illegible] that other variants are clinically actionable

Only clinically actionable somatic variants ad VUSs that are biologically relevant are reported: A complete list of variants is [illegible] upon request Germline VUSs are reported only for 54 well-established cancer predisposition genes or those with sufficient clinical evidence for an [illegible] paediatric cancer and are marked with an utn in Appendix 1, Variant classification, particularly for VUSs, may change as more information becomes [illegible] as inheritance and allelic infomation is obtained by familial segregation studies. When parental segregation studies are performed, accurate assignm[illegible] family [illegible] is critical to the interpretation of test results; therefore, a separate test to confim the family relationships may be performed. The mode of [illegible] for each gene is obtained from OMIM; when available. If not available in OMIM; the mode of inheritance is obtained from the scientific literature [illegible] included in the report.

Variants classified as benign or likely benign are not reported Synonymous variants, 3' and 5' UTR var[illegible] and intronic variants (beyond +10 base pairs) are not reported unless known to be pathogenic or predicted to disrupt splicing as indicated by the follo[illegible] programs SpliceSiteFinder, MaxEntScan, NNSPLICE , GeneSplice or predicted to disrupt translation initiation.

Genetic variants not related to the primary reason for referral, also described by the Am[illegible] College [illegible] Medical Genetics guidelines (Miller et al (2021) PMID: 34012066) , may be identified by this assay. Variants in these genes that are identified [illegible] are included on this report if they have clinical utility in the context of an increased heritable risk of cancer Pathogenic germline variants in non-c[illegible]cer [illegible] are considered incidental findings and will not be included in the oncology report; unless consented to by the patient VUSs in non-cancer disorders w[illegible] be re[illegible]rted.

**ANALYTICAL AND CLINICAL SENSITIVITY:**

Next-Generation sequencing may not detect all sequence variants associate[illegible] with the [illegible]tient's cancer. For gemline samples, this test is >95% sensitive for detecting substitution variants, >90% sensitive for detecting small insertions [illegible] [illegible]% sensitive for detecting CN changes For tumour samples , this test is >90% sensitive for detecting substitution variants and >80% sensitive for [illegible] [illegible]rtions or deletions. The threshold for reporting clinically actionable variants from fresh-frozen samples is a VAF of 5% for SNVs and a VAF of 10% [illegible] [illegible] deletions. For FFPE samples, the threshold for reporting clinically actionable variants is a VAF of 1% for SNVs and small insertions and deletions. Clini[illegible] actionable variants detected below these VAFs may also be included o this report; however; they are below the validated thresholds for this test and are [illegible] by an orthogonal method For fresh-frozen samples, this test is >90% sensitive for detecting focal (<5 Mb) on-target copy number [illegible] [illegible] >95% sensitive for detecting large segmental (>5 Mb), whole chromosome arm, whole chromosome CN variants and regions of copy neutral LOH (710 M[illegible] The limit-of- detection for CN variants by this test is 300-400 bp in size for gains/losses encompassing on-target regions. Tumour samples with at least 25% tumour [illegible] required for reliable detection of CN changes. Somatic variants that occur at low levels in the tumour tissue because of tumour heterogeneity or low [illegible] [illegible]lularity may not be detected Results must be interpreted in the context of tumour cellularity when available. Complex small insertions deletions or CN [illegible] in clini[illegible] actionable genes detected by the NGS assay may be further verified by Sanger sequencing or QPCR.

Regions beyond +10 base pairs of the splic[illegible] [illegible] not consistently covered as such, this test does not provide information about non-coding regions of the gene (e g regulatory 5' and 3' UTR[illegible] [illegible]ntronic regions) unless specifically indicated The failure to detect a causative genotype by this assay does not exclude the possibility of ano[illegible]er mole[illegible]lar basis for the patient's cancer NGS technologies capture techniques and associated bioinformatics analysis have limitations ad may resu[illegible] failure [illegible] detect s[illegible]e sequence variants Highly homologous and pseudogene sequences, as well as GC-rich and repetitive regions, may interfere with accurate d[illegible] of [illegible] [illegible] sequencing analyses. A subset of exons is not included in this analysis due to technical lirtations of this assay. This list is available upon request. [illegible] typ[illegible] genomic alterations known to cause disease will not be detected by this NGS assay including, but not limited to, structural rearrangement[illegible] (e.g balan[illegible] [illegible]locations, inversions and repeat expansions) of the targeted exons.

This test was dev[illegible] [illegible] performance characteristics determined by the Chimwald Molecular Diagnostics Lab as required by IQMH and CLIA '88 regulations This laboratory has [illegible] and verified the test's accuracy and precision The test has not been cleared or approved by Health Canada or the US Food and Drug Administration (FDA) H[illegible]th Canada and the FDA have determined that such clearance or approval is not necessary.

#### Appendix 1: 13 genes included on the Comprehensive Cancer Panel.

EGFR NOTCH4 MBD1 NTRK1 HOXA9 HMGA2 HIST1H4D NFIB POU5F1 RAF1 FLI1 FUBP1 ATIC

Mock Specimen

Eagledwell Health Complex
EAGLEDWELL HEALTH ALLIANCE

**Laboratory Medicine Division**

123 Sample St, Eagledwell AB, A1B 2C3

| | |
|---|---|
| Name: | **Redacted** |
| Client: | Pediatric Hospital of Annsburg (Annsburg) |
| PIN/MRN: | **Redacted** |
| Ref #: | Redacted |
| Visit #: | Redacted |
| Location: | - |
| Sex: | Male |
| DOB: | Redacted Age: Redacted |
| Ordering Dr: | Redacted |
| Attending Dr: | Redacted |

**Molecular Genetics**

#### Familial Adenomatous Polyposis (FAP)
#### Extra-Intestinal Manifestations (CHRPE, CMV Thyroid, Desmoid) Screen Report

**Sample ID**: MD-24-0004213 / PAO0123456 **Specimen**: Amplified DNA **Received**: [illegible]22-05-[illegible] 19:05:36

| Variant* | Classification |
|---|---|
| DYNC1H1:c.12700T>G , p.Ser4234Ala + | Variant of uncertain clinical significance |

**Interpretation:**

A homozygous variant, NM_001376.5(DYNC1H1):c.12700T>G, p.Ser42[illegible]Ala was detected in exon 19 of this gene. The allele frequency of this variant in the population databases is: gnomAD: (0.0438% overall). The classification of this variant in databases with clinically curated data is: ClinVar: "Variant of uncertain clinical significance". In silico [illegible] the effect of this amino acid change on protein structure and function is: SIFT: "tolerated"; PolyPhen-2: "neutral"; Align-GVGD: "Class [illegible]5". This variant has not been reported in the literature. Based on the current evidence, we interpret this variant as variant of uncertain clinical significance (ACMG category).
Genetic counseling and clinical following are recommended.

**Disease Characteristics:** Familial Adenomatous Polyposis (FAP) is a colon cancer predisposition syndrome associated with development of thousands of adenomatous colonic polyps in early years of life. Extracolonic manifestations associated with this syndrome are variably present and include: polyps of the gastric fund[illegible]denum, osteomas, dental anomalies, congenital hypertrophy of the retinal pigment epithelium (CHRPE), soft tissue tumors, de[illegible]oid tumors, and associated cancers (PMID: 18612695). Germline mutations in APC gene have been associated with autosomal dominant form of FAP and germline mutations in MUTYH gene have been linked with autosomal recessive form of this disease. This genetic test may confirm a diagnosis and help guide treatment and management decisions.

**Methodology:** All coding exons, [illegible]0 bp of flanking intronic sequence and the regions listed below the Genes Tested table are enriched using an EHC custom targeted hy[illegible]ocol (Roche Nimblegen), followed by high throughput sequencing (Illumina). Sequence variants and copy number changes are assessed and interpreted using clinically validated algorithms and commercial software (SoftGenetics: Nextgene, Geneticist Assistant, Mutation Surveyor; and Alamut Visual). All exons have >300x mean read depth coverage, with a minimum 100x coverage at a single nucleotide resolution. This assay meets the sensitivity and specificity of combined Sanger sequencing and MLPA copy number analysis. Variants interpreted as either ACMG category 1, 2 or 3 (pathogenic, likely pathogenic, VUS; PMID: 25741868) are confirmed using Sanger sequencing, MLPA, or other assays when necessary. ACMG category 4 and 5 variants (likely benign, benign) are not reported, but are available upon request. This assay has been validated at a level of sensitivity equivalent to the Sanger sequencing and standard copy number analysis (>99%; PMID: 27376475, 28818680). Sample fidelity is assessed by matching sequencing results with the Exome QC genotyping assay (Agena).

**Disclaimer:** This test was developed and its performance determined by this laboratory, which is accredited by Accreditation Canada. Utilization of the reported results for clinical purposes remains the full responsibility of the practitioner. Testing is highly accurate, however, rare diagnostic errors can arise from specimen mishandling or misinterpretation, and it has been reported that as much as 0.5% error rate may occur in any of the pre-analytical/analytical or post analytical phases of the test (PMID:11978595).

**Genes Tested** (hg19;HGVS nomenclature):
HERPUD1, NCAPD3, SIN3A, LRRK1, BTK, EPHB1, IDH2, LMO2, ACSL3, KDM4C, FANCA, CSF2RA, FUBP1, CAMTA1, LPP, TPO, DYNC1H1,

Mock Specimen

**Intronic Regions Tested** (in addition to 20bp flanking coding exons):
NM_001127511.2(APC):c.-192A>G NM_001127511.2(APC):c-191T>C NM_001127511.2(APC):c.-190G>A NM_001128425.1(MUTYH):c.504+19_5

(Electronically signed by)
Redacted
2022-05-12 09:31:15

---

Printed: 2022-05-12 09:31:15
Request ID: Redacted

Mock specimen

Hedgefield
Health Center
We Care

| | | | |
|---|---|---|---|
| **Patient:** | **Redacted** | | |
| **D.O.B.:** | Redacted | **Legal Sex:** | Redacted |
| **MRN:** | Redacted | **Family Doctor:** | Redacted |
| **Account #:** | Redacted | **Registration Date:** | Redacted |
| **Health Card #:** | Redacted | **Specimen #:** | Redacted |

**Specimen Details**

| | Collected: | Received: | Type: | Source: |
|---|---|---|---|---|
| **Redacted** | 2022-10-09 22:52:31 | 2022-10-09 23:59:06 | Blood | Blood, Venous |

**Hereditary Cancer Panel** (Final result)

#### Referring Diagnosis:

Hereditary Breast/Ovarian/Prostate Cancer

#### Test Summary

| Panel | Platform | Software | Genome Build |
|---|---|---|---|
| SOPHiA Custom Hereditary Cancer Solution (CHCS_AA_VI) | Illumina NextSeq 550 | Illumina LRM v.2.4, SOPHiA DDM v.5.10 | GRCh37 |

#### Interpretation

**RESULT: A pathogenic germline variant was detected in the BCL9 gene.**

| Classification | Accession(Gene):cDNA,Protein | Zygosity |
|---|---|---|
| Likely pathogenic | NM_004326.4(BCL9):c.356A>C, p.Asp119Ala | heterozygous |

**Interpretation**: A pathogenic variant c.356A>C , p.Asp119Ala , was detected in the BCL9 gene. This missense variant is predicted to result in amino acid substitution of aspartate at codon 119 with alanine . this variant is present at a low frequency in the general population in the Genome Aggregation Database (gnomAD ALL: 0.03710935 %, dbSNP: rs496517823, ACMGG PM2), as well as in unaffected individuals [PMID: 20352372 ]. Although the majority of pathogenic variants in are truncating, this variant has been reported in individuals with breast cancer or ovarian cancer [PMID: 26030354 ] (ACMGG PS4_Supporting) In silico analyses are concordant regarding the pathogenic effect this varinat may have on protein structure and function. (ACMGG BP4), however, functional analyses have not been performed to verify these predictions. Clinvar contains an entry for this variant (Variation ID: 934618 ) where it is classified as likely pathogenic by the majority of submitting laboratories. Given the available evidence, our laboratory currently classifies this variant as likely pathogenic (ACMGG: PS4_Supporting, PMZ, BPI, BPA).

**Recommendations**: Medical screening and management should rely on clinical findings and family history. An explanation of these results is available through consultation with the local Genetics clinic.

**Please note**: Variants determined to be benign or likely benign are not included on this report; but are available upon request.

**HBOP1 Genes examined**: EPHA6, IDH2, RANBP2, CTTN, TAF15, TSC2, ZMYM4, BCL9, PKHD1, MAFB, BRCA2, EPHA2,

Mock Specimen

Hedgefield General,
123 Sample St,
Hedgefield, AB, A1B 2C3,
T: (123) 456-7890

SH3GL1, CBFB,

**Methodology**: Massively parallel sequencing of coding exons and exon-intron junctions (+/-20 bp) of the above genes was performed with the Sophia CHCS_AA_v1 panel; sequence variant analysis and duplication/deletion analysis using sequence depth assessment was performed on the Sophia DDM software: Confirmation of copy number variants with an alternate method (MLPA or microarray) was performed when necessary and possible.

**Variant Classification Rules**: ACMG Guidelines. Richards et al, Genet Med. 17:405-424

**Nomenclature Guidelines**: HGVS Guidelines, varnomen.hgvs.org, Version 20.05.

**Limitations**: The following may not be detected: low-level mosaics (< 20% allele frequency), regulatory and deep intronic regions; breakpoints of large deletions or duplications. Diagnostic errors may occur due to rare sequence variants, inaccurate family histories or sample mixup.

#### Signed By

Redacted on 2022-10-11 03:35:44

Blood specimen REDACTED from Blood, Venous Unspecified. Ordered by Redacted. Authorized by Redacted. Collected: 2022-10-09 22:52:31, Received: 2022-10-09 23:59:06. Verified: 2022-10-11 03:35:44. Results by Hedgefield Health Centre Molecular Genetics Lab.

#### Resulting Labs

**Hedgefield General Hospital Laboratory - Molecular Genetics** MOLGEN LAB, 123 Sample St.; Hedgefield A1B, 2C3

**Lab Section:** Genetics

**** END OF REPORT ****

If you have received this report in [illegible] by Fax to Health Information Management at Hedgefield Health Centre at 123-456-7899. If you don't have access to fax, or have other questions, please call 123-456-7890 ext. 0001

Mock Specimen

**Toronto Cancer Hospital**

Division of Tumor Sequencing and Diagnostics

123 Main St. West,
Toronto, ON, A4B5G9

### Draft PATHOLOGY LABORATORY RESULTS

Mock specimen

#### Patient Info:

Name:
DOB:
Sex:

#### Patient Information:

Physician Name:
Health Card:
MRN #:
Clinic: Bea[illegible]ngton Health Centre (Be[illegible]rli[illegible]ton)

| Procedure Date | Accession Date | Report Date |
|---|---|---|
| 2022-10-12 06:03:18 | 2022-10-14 00:22:41 | 2022-10-15 13:32:59 |

#### Report Details

Genome Reference - GRCh3[illegible]

Sequencing Information - Targeted variant testing, Karyotyping

Referral Reason - Research

Analysis Location - Toronto Cancer Hospital (Toronto)

### Table of findings:

| Gene | Information | Interpretation |
|---|---|---|
| [illegible]ST[illegible]H1B | 3 c.181T>A p.Ser61Thr heterozygous | Likely pathogenic |

**Summary:** A likely pathogenic variant detected.. The details regarding the specific mutations are included below.

**Toronto Cancer Hospital**

Division of Tumor Sequencing and Diagnostics

123 Main St. West,
Toronto, ON, A4B5G9

### Variant Interpretation:

The interpretation of these variants is as follows: A likely pathogenic variant was detected in the sample.

##### Variant 1 of 1

| Gene | Variant | Amino | Zygosity |
|---|---|---|---|
| HIST1H1B | c.181T>A | p.Ser61Thr | heterozygous |

###### Likely pathogenic

The g.27834750T>A variant occurs in chromosome chr6 , within the HIST1H1B gene, and it causes c.181T>A change at position 61 in exon 3 , causing the mutation p.Ser61Thr . This mutation has been identified in 45 families. It has a population frequency of 1.99e-03 (85[illegible] alleles in 428164 total alleles tested), indicating it is a uncommon variant in the general population. It

ClinVar and other genomic databases report the HIST1H1B c.181T>A variant as clinically relevant based on aggregated evidence.

This variant is classified as likely pathogenic. It is believed to negatively impact protein function and may play a role in disease development in affected individuals.

The affected nucleotide lies within a region that is highly conserved across vertebrate species, which suggests functional importance and evolutionary constraint.

This variant is implicated in oncogenesis and other disease processes according to ClinVar records (VCV accession: VCV006458160).

Supporting studies and case reports can be found in the scientific literature. Relevant PubMed references include: 287134430, 314433523, 335220977, 923644379, 342011708 .

Mock Specimen

#### Toronto Cancer Hospital

Division of Tumor Sequencing and Diagnostics

123 Main St. West,
Toronto, ON, A4B5G9

In accordance with existing evidence, this variant is therefore classified as a likely pathogenic variant.

### Test Details:

**Genes Analyzed:** HIST2H4A. SMAD4. RIMS2. GATA1. BCOR. DACH2. NACA. ACSL6. PTCH1. CDK6. HIST1H1B. MPO. WHSC1. 13 total genes tested.

**mRNA ref sequences tested NM_:** NM_003548.2 NM_005359.6 NM_0013[illegible]8484.3 NM_002049.4 NM_001123385.2 NM_053281.3 NM_001365896.[illegible] NM_001009185.3 NM_000264.5 NM_001145306.2 NM_005322.2 NM_000250.[illegible] NM_1333[illegible]0.2

##### Recommendations

A precision oncology approach is advised. These mutations are known oncogenic drivers, linked to constitutive pathway activation. Targeted therapies, including hormone-correcting agents, may be considered, guided by clinical judgment. PI3K inhibitors could be explored in trials for PIK3CA-mutated cases. Germline testing is not indicated, as all mutations are consistent with somatic events. A multidisciplinary tumour board review is recommended to integrate findings into care. Additional testing may be pursued at the physician's discretion.

##### Methodology

ctDNA was sequenced with Targeted variant testing and was analyzed using Karyotyping covering all coding exons and adjacent intronic regions. Target enrichment was performed with hybrid capture (Twist Bioscience), followed by Illumina NextSeq sequencing. Reads were aligned to GRCh37 using BWA-MEM, and variants called with GATK. Annotation was performed in VarSeq using population databases, predictive algorithms, and ClinVar. CNVs were assessed with CNVkit and confirmed by MLPA when applicable. Regions with pseudogene interference, such as PMS2, were validated using long-range PCR and Sanger sequencing. Mean read depth exceeded 300x, with a minimum threshold of 50x. Analytical sensitivity is >99% for SNVs/indels and >95% for exon-level CNVs. **Only variants classified as pathogenic, likely pathogenic, or variants of uncertain significance (VUS) are reported**, per ACMG/AMP guidelines (PMID: 25741868).

##### Limitations

This test was developed and validated in a certified clinical laboratory. Limitations include reduced sensitivity in pseudogene regions (e.g., PMS2, CHEK2), and inability to detect certain structural variants (e.g., MSH2 inversions), deep intronic changes, or low-level mosaicism. PMS2 exons 11–15 are not analyzed. Interpretations reflect current knowledge and may be updated as new evidence emerges.

Mock Specimen

**Toronto Cancer Hospital**
Division of Tumor Sequencing and Diagnostics

123 Main St. West,
Toronto, ON, A4B5G9


### Report Electronically Verified and Signed by:

Mock specimen

### FAWNGROVE GENERAL HOSPITAL

Molecular Genetics
Laboratory
123 Sample St., Fawngrove
AB A1B 2C3
T 123.456.7890
F 123.456.7899
fgh.ca

#### MOLECULAR GENETICS REPORT

Dr. REDACTED
Lionsburgh Medical Centre (Lionsburgh)
Address Redacted
Address Redacted

**Last name:** Redacted
**First name:** Redacted
**DOB:** Redacted
**Sex:** REDACTED
**HC#:** Redacted

---

| | | |
|---|---|---|
| **Lab #:** JL2O07143B | **Date Received:** 2023-05-12 22:55:49 | **Reason for** [illegible]**:** |
| **Sample Type:** Blood | **Date Requested:** 2023-05-10 19:30:30 | Heredita[illegible]y [illegible] & Ovarian Cancer Testing |
| **Genetic Clinic #:** - | **Date of Report:** 2023-05-14 23:49:21 | [illegible] Gene panel |

---

| **Summary:** | **Gene** | **Variant** | **Clinical Significance** |
|---|---|---|---|
| | PMS2 | c.103C>G (p[illegible]) | Variant of uncertain clinical significance |

---

**Interpretation:**

A Variant of uncertain clinical significance i[illegible] 7 of t[illegible] PMS2 gene was detected in this individual. This variant, c.103C>G , p.Leu35Val , is predicted to [illegible] in the substitution of valine for the leucine at codon 35 . This variant has been reported [illegible] Clir[illegible]var by multiple submitters as Variant of uncertain clinical significance . It does not appear to have been dis[illegible] the literature. The allele frequency is 1949 / 494366 in the Genome Aggregation Database (v2.1.1[illegible]). In silico anlyses (Align GVGD, MutationTaster; POLYPHEN-2 and SIFT) were consistent i[illegible] predicti[illegible]g this variant to be variant of uncertain clinical significance . This nucleotide is not conser[illegible] = 0.1). This variant would be classified as variant of uncertain clinical significance .

Family studies [illegible] be [illegible] in further elucidating the significance of these variants with respect to familial breast/ovarian [illegible] Genetic counseling and clinical follow-up are recommended.

**Analysis:**

Genes analyzed [illegible] genes): DEPDC5 (NM_001242896.3), CDKN2A (NM_000077.5), PMS2 (NM_000535.7), TCF7 (NM_003202.5), NKX2-1 (NM_001079668.3), ATP1A1 (NM_000701.8),

Genomic DNA was enriched for the coding regions of genes listed above by a hybridization-based capture method using TruSight Rapid Capture/Canadian Consortia Inherited Cancer Panel kit (Illumina). The enriched libraries were sequenced the NextSeq 500 System (Illumina) then aligned tp the entire GRCh37 (hg19) human reference genome and variants were called and annotated using NextGENe (v2.4.2.3) and Geneticist Assistant (v1.4.2) softwares (Softgenetics). The description of the genetic variants observed follows the HGVS guidelines on nomenclature [Hum Mutat (2016) 37.564-569] and the American College of Medical Genetics recommendations for standards for interpretation of sequence Variations [Genet Med (2015) 17.405-424; Am J Hum Genet (2016) 98.801-817]. Only variants found in the coding regions and 15 nucleotides into the introns, as well as into the 5'- and 3'-UTR regions of these genes, are reported. With the exception of the 5' end of MLH1 exon 12 where intronic basepairs are analyzed. All sequence variants interpreted as pathogenic, likely pathogenic or uncertain significance were re-analyzed using Sanger sequencing when internal laboratory quality

Mock specimen

### FAWNGROVE GENERAL HOSPITAL

Molecular Genetics
Laboratory
123 Sample St., Fawngrove
AB A1B 2C3
T 123.456.7890
F 123.456.7899
fgh.ca

metrics were not met. Low coverage regions containing at least one nucleotide with a read depth lower than 20 were re-analyzed using Sanger sequencing. Pathogenic, likely pathogenic and uncertain variants in PMS2 and CHEK2 (exons 11-15) were confirmed by long range PCR of the PMS2 and CHEK2 genes, respectively, to exclude the possibility of the variants being present in the pseudogenes of PMS2 and CHEK2. Exonic copy number changes were analyzed by the RPKM read counting method (reads per kilobase per million mapped reads) using NextGENe software [Bioinformatics (2012) 28.2747-2754]. All deletions/duplications detected were confirmed by MLPA analysis (MRC-Holland). Variants are listed as heterozygous unless otherwise indicated. Benign and likely benign variants are not listed but are available upon request.* For EPCAM, only copy number analysis is performed.

Limitations: This test does not detect low level mosaicism, large duplications and deletions or structural rearrangements: Variants in PMS2 may not be detected due to interference with the presence of pseudogenes. Sequence variants in exon 15 of PMS2 are not analyzed unless immunohistochemistry indicated negative for PMS2. Variants in regions containing long stretches of homopolymers may not be detected.

Test sensitivity: ~99%, Test specificity: ~100%

Disclaimers: This test is based on our current understanding of the molecular knowledge of hereditary breast/ovarian cancer. There is a possibility that clinically significant changes in variant classification will occur through new discovery, enhanced clinical correlation and functional or epidemiologic studies. This test was developed and its performance characteristics were determined by the Molecular Genetics Laboratory at Fawngrove General Hospital, which is accredited under the Institute for Quality Management in Healthcare (IQMH).

Mock specimen

Signed: Redacted
Laboratory Geneticist

**Golden Field Health | Girton General**
GFH Health Nexus

**Molecular Genetics Laboratory**
123 Sample St. Girton, AB, A1B 2C3
t: 123-456-7890 x4457 f: 123-456-7899

| | | | |
|---|---|---|---|
| Clinic ID: | | Medical Record #: | Redacted |
| Clinic Name: | Narwald Central Hospital (Narwald) | Last Name: | Redacted |
| Physician: | Redacted | First, Middle: | Redacted |
| Procedure Date: | 2025-12-03 22:04:40 | DOB/Sex: | Redacted/Redacted |
| Accession Date: | 2025-12-05 18:15:33 | Health Card #: | Redacted |
| Report Date: | 2025-12-06 15:27:04 | Visit #: | Redacted |

### FINAL LABORATORY GENETICS REPORT: GEN-24-XXXX

Copies to: REDACTED

**Molecular Lab#:** M24-XXXX

**SPECIMEN:** Blood

**TEST DESCRIPTION:** Targeted variant testing

**TEST INDICATION:** Personal history of VHL; Positive family history

**RESULT:** POSITIVE

**DNA FINDING:**

| Gene, Transcript | Variant, Prediction | Zygosity | Interpretation |
|---|---|---|---|
| *NTRK3*, NM_001012338.3 | c.2297A>C, p.Tyr766Ser | heterozygous | Variant of uncertain clinical significance |
| *CHEK2*, NM_007194.4 | c.1629G>A, p.Leu543= | heterozygous | Variant of uncertain clinical significance |
| *NTRK3*, NM_001012338.3 | c.1650A>T, p.Gly550= | heterozygous | Variant of uncertain clinical significance |
| *NTRK3*, NM_001012338.3 | c.1404G>A, p.Val468= | heterozygous | Likely pathogenic |
| *ETV1*, NM_004956.5 | c.995G>T, p.Arg332Leu | heterozygous | Likely pathogenic |

**INTERPRETATION:** Sequencing identified two likely pathogenic variants and three variants of uncertain clinical significance listed above. The result is consistent with clinical history.

**RECOMMENDATION:** Genetic counselling is recommended for this individual and the family. Targeted variant testing is available for other family members. Clinical management strategies may be recommended according to established guidelines. To check on the status of a variant or for assistance in locating nearby genetic counselling services please contact Advanced Molecular Diagnostics by email at Redacted.

**VARIANT INFORMATION:**

NTRK3, EXON[illegible], c.2297A>C, p.Tyr766Ser, Heterozygous, Variant of uncertain clinical significance

The NTRK3 c.2297A>C, p.Tyr766Ser variant was identified in multiple individuals with cancer syndromes ( PMID:34681835, PMID:36819219 ). The variant was also identified in ClinVar (classified as Variant of uncertain clinical significance by multiple submitters). The variant was identified in controls in 335 of 669,189 chromosomes at a frequency of 0.000500606 (Genome Aggregation Database Nov 1 2023 v4.0.0). Functional studies did not demonstrate a disruption of protein stability and function ( PMID:14452504, PMID:20644767 ). The residue is moderately conserved across mammals and other organisms, and computational analyses (PolyPhen-2, SIFT, AlignGVGD, MutationTaster) suggest that the variant may not impact the protein. The variant occurs outside of the splicing consensus sequence and in silico or computational prediction software programs (SpliceSiteFinder, MaxEntScan, NNSPLICE, GeneSplicer) do not predict a difference in splicing. In summary, based on the above information, the clinical significance of this variant is classified as Variant of uncertain clinical significance.

**References (PMIDs):** 38437232, 19393714, 32965301, 33647615, 22705651, 17359370, 10003384, 33138955, 39681475, 13934487

CHEK2, EXON04, c.1629G>A, p.Leu543=, Heterozygous, Variant of uncertain clinical significance

Mock Specimen

The CHEK2 c.1629G>A , p.Leu543= variant was identified in multiple individuals with cancer syndromes ( PMID:36203817, PMID:21350524, PMID:26520790, PMID:37651900, PMID:35795346, PMID:37620357, PMID:34819921, PMID:18807746, PMID:23267995, PMID:31701258 ). The variant was also identified in ClinVar (classified as Variant of uncertain clinical significance by multiple submitters). The variant was identified in controls in 0 of 308,412 chromosomes at a frequency of 0 (Genome Aggregation Database Nov 1 2023 v4.0.0). Functional studies did not demonstrate a disruption of protein stability and function ( PMID:33443251, PMID:36503389 ). The residue is moderately conserved across mammals and other organisms, and computational analyses (PolyPhen-2, SIFT, AlignGVGD, MutationTaster) suggest that the variant may not impact the protein. The variant occurs outside of the splicing consensus sequence and in silico or computational prediction software programs (SpliceSiteFinder, MaxEntScan, NNSPLICE, GeneSplicer) do not predict a difference in splicing. In summary, based on the above information, the clinical significance of this variant is classified as Variant of uncertain clinical significance.

**References (PMIDs)**: 39462422, 12394032, 33626411, 16717469, 36222775, 19500981, 38805399, 34663955, 14592163

---

NTRK3, EXON15, c.1650A>T, p.Gly550=, Heterozygous, Variant of uncertain clinical significance

---

The NTRK3 c.1650A>T , p.Gly550= variant was identified in multiple individuals with cancer syndromes ( PMID:37643043, PMID:10915644, PMID:37357182, PMID:10238270, PMID:37241778, PMID:37905042, PMID:[illegible] ). The variant was also identified in ClinVar (classified as Variant of uncertain clinical significance by multiple submitters). The variant was identified in controls in 749 of 625,270 chromosomes at a frequency of 0.001197883 (Genome Aggregation Database Nov 1 2023 v4.0.0). Functional studies did not demonstrate a disruption of protein stability and function ( PMID:[illegible]87571, PMID:26918322 ). The residue is moderately conserved across mammals and other organisms, and computational analyses (PolyPhen-2, SIFT, AlignGVGD, MutationTaster) suggest that the variant may not impact the protein. The variant occurs outside of the splicing consensus sequence and in silico or computational prediction software programs (SpliceSiteFinder, MaxEntScan, NNSPLICE, GeneSplicer) do not predict a difference in splicing. In summary, based on the above information, the clinical significance of this variant is classified as Variant of uncertain clinical significance.

**References (PMIDs)**: 31101344, 27601627, 27250168

---

NTRK3, EXON14, c.1404G>A, p.Val468=, Heterozygous, Likely pathogenic

---

The NTRK3 c.1404G>A , p.Val468= variant was identified in multiple individuals with cancer syndromes ( PMID:11202931, PMID:20932361, PMID:39856601, PMID:28052840, PMID:[illegible]795, PMID:28402606, PMID:26642082 ). The variant was also identified in ClinVar (classified as Likely pathogenic by multiple submitters). The variant was identified in controls in 0 of 346,741 chromosomes at a frequency of 0 (Genome Aggregation Database Nov 1 2023 v4.0.0). The variant has been found in the literature to segregate with disease in multiple families ( PMID:33601685, PMID:14534629, PMID:19433327 ). Furthermore, in functional studies, the variant is demonstrated to disrupt protein stability and function ( PMID:34168753, PMID:10894554 ). The residue is conserved across mammals and other organisms, and computational analyses (PolyPhen-2, SIFT, AlignGVGD, MutationTaster) suggest that the variant may impact the protein. The variant occurs outside of the splicing consensus sequence and in silico or computational prediction software programs (SpliceSiteFinder, MaxEntScan, NNSPLICE, GeneSplicer) do not predict a difference in splicing. In summary, based on the above information, the clinical significance of this variant is classified as Likely pathogenic.

**References (PMIDs)**: [illegible], [illegible]472434, 12662861, 31111488, 21012642, 36958182, 36103719

---

ETV1, EXON03, [illegible] p.Arg332Leu, Heterozygous, Likely pathogenic

---

The ETV1 c.995G>T , p.Arg332Leu variant was identified in multiple individuals with cancer syndromes ( PMID:37857858, PMID:24512206, PMID:24815009, PMID:19869115, PMID:19473753 ). The variant was also identified in ClinVar (classified as Likely pathogenic by multiple submitters). The variant was identified in controls in 0 of 615,731 chromosomes at a frequency of 0 (Genome Aggregation Database Nov 1 2023 v4.0.0). The variant has been found in the literature to segregate with disease in multiple families ( PMID:27954959, PMID:10651842, PMID:19307631, PMID:23057412, PMID:22389090, PMID:29139819, PMID:19225244 ). Furthermore, in functional studies, the variant is demonstrated to disrupt protein stability and function ( PMID:22050031, PMID:16876650 ). The residue is conserved across mammals and other organisms, and computational analyses (PolyPhen-2, SIFT, AlignGVGD, MutationTaster) suggest that the variant may impact the protein. The variant occurs outside of the splicing consensus sequence and in silico or computational prediction software programs (SpliceSiteFinder, MaxEntScan, NNSPLICE, GeneSplicer) do not predict a difference in splicing. In summary, based on the above information, the clinical significance of this variant is classified as Likely pathogenic.

**References (PMIDs)**: 22292790, 21439022, 13358876, 20279152, 23840069, 18642243, 37997024, 10935016, 20411437, 11042058

Mock Specimen

**Assessment Date**: 2025-12-08 15:27:04

---

**BACKGROUND INFORMATION** : The VHL gene is associated with the autosomal dominant cancer syndrome von Hippel Lindau disease, which is characterized by retinal hemangiomas, cerebellar and spinal hemangioblastomas, renal cell carcinoma and pheochromocytomas. VHL is often separated into VHL type 1, which has low risk for pheochromocytomas, and VHL type 2, which has a high risk for pheochromocytomas. In general, penetrance is high and age-dependent, with nearly complete penetrance by age 65. Autosomal dominant conditions have a 50% chance of transmission between first degree relatives: Not all cases will have a variant detectable by this test; therefore, a negative result does not rule out von Hippel Lindau disease in a family. The classification of DNA variants may change over time as new information becomes available. The significance of a DNA alteration should always be interpreted in the context of the individuals clinical manifestations.

**TESTING METHODOLOGIES** : Gene sequencing and exon-level copy number variant (CNV) detection were performed by Next-Generation Sequencing (NGS) on an Illumina NovaSeq 6000 or NextSeq 550 Instrument. Following enrichment using the Illumina DNA Prep with Enrichment for the full coding regions and splice sites (+ or - 15 base pairs from the exon boundaries) as well as targeted non-coding variants (list available upon request), with a minimum coverage of [illegible]x for the genes listed in the Test Description (reference sequences can be found at: https://goldenfieldhealth.ca/forms/[illegible]/, version 2023.01). Regions with coverage less than 20x are covered by Sanger sequencing: In addition, the DNA was analyzed for the presence of the targeted variant: NM_000551.3(VHL):R167W. DNA variant positions are provided using HGVS nomenclature. Low confidence variants (e.g. low alternate allele frequencies, pseudogene regions, and high GC content) and copy number variants are confirmed by Sanger sequencing (with long-range PCR for certain pseudogene regions) and/or multiplex ligation-dependent probe amplification (MLPA) analysis on a 3730xl DNA Analyzer following an independent isolation of DNA from blood, if available. Variants outside the regions of interest may not be detected or analyzed. Benign or likely benign variants are not included on this report, but are available upon request. Sequencing may not detect all variant types including variants in coding or non-coding regions; including but not limited to variants that could affect gene expression or splicing, deletions, duplications, insertions, indels, chromosomal aberrations or rearrangements. This test has an analytical sensitivity and specificity of >99% for DNA substitutions and small deletions or duplications (up to 5bp) as well as exon-level or full gene deletions or duplications.

The detection of single nucleotide variations was performed following the Best Practices for Variant Calling with the Genome Analysis Toolkit (GATK 4.1.0) published by the Broad Institute. DNA sequence mapping is performed using the GRCh37/hg19 assembly as the genome reference build. Detection of exon-level copy number variations uses the published and internally validated tool ExomeDepth 1.1.0.

These test results presuppose that the sample received by the Advanced Molecular Diagnostics Laboratory for testing was in fact from this patient and was not contaminated nor subject to sample mix-Up. Results do not include the possibility of sample mix-Up or laboratory error, which is repo[illegible] <1%. Rare diagnostic errors can result from incorrectly assigned family relationships (e.g. non-paternity) or DNA alterations (e.g. DNA variant under a primer or probe binding site; the presence of pseudogene artifacts). DNA variants or alterations outside the region of analysis will not be detected: This test was not designed to detect somatic variant alterations. These tests were developed and their performance characteristics determined by Golden Field Health, Molecular Genetics Laboratory. This laboratory is accredited to ISO 15189 Plus by Accreditation Canada Diagnostics. These tests were validated according to accepted practice guidelines for clinical molecular genetic testing by the ACMG and CCMG.

Report Electronically Signed by:
Redacted
Laboratory Director

### FAWNGROVE GENERAL HOSPITAL

Molecular Genetics
Laboratory
123 Sample St., Fawngrove
AB A1B 2C3
T 123.456.7890
F 123.456.7899
fgh.ca

#### MOLECULAR GENETICS REPORT

Dr. REDACTED
King's Health Campus (Klagenbeck)
Address Redacted
Address Redacted

**Last name:** Redacted
**First name:** Redacted
**DOB:** Redacted
**Sex:** REDACTED
**HC#:** Redacted

---

**Lab #:** JL2O07143B
**Sample Type:** Blood
**Genetic Clinic #:** -

**Date Received:** 2025-11-06 03:35:06
**Date Requested:** 2025-11-04 05:46:59
**Date of Report:** 2025-11-07 01:55:30

**Reason for referral:**
Hereditary Breast & Ovarian Cancer Testing
**Test Requested:** Targeted variant testing

---

**Summary:**

| Gene | Variant | Clinical Significance |
|---|---|---|
| DDX5 | c.1144T>G (p.[illegible]) | Likely pathogenic |
| KDM5A | c.674C>A (p.[illegible]225[illegible]er) | Likely pathogenic |
| COL11A1 | c.3264C[illegible]A ([illegible]1088=) | Variant of uncertain clinical significance |

---

**Interpretation:**

A Likely pathogenic in exon 6 of the DDX5 gene was detected in this individual. This variant, c.1144T>G , p.Trp382Gly , is predicted to result in the substitution of glycine for the tryptophane at codon 382 . This variant has been reported in Clinvar by multiple submitters as Likely pathogenic . It does not appear to have been discussed in the literature. The allele frequency is 756 / 655697 in the Genome Aggregation Database (v2.1.1, non-cancer). In [illegible] (Align GVGD, MutationTaster; POLYPHEN-2 and SIFT) were consistent in predicting this variant to be likely pathogenic . This nucleotide is conserved (phyloP = 0.7). This variant would be classified as likely pathogenic .

A Likely pathogenic in exon 15 of the KDM5A gene was detected in this individual. This variant, c.674C>A , p.Ser225[illegible] , is predicted to result in an early translation termination at codon 225 . This variant has been reported in Clinvar by multiple submitters as Likely pathogenic . It does not appear to have been discussed in the literature. The allele frequency is 571 / 494115 in the Genome Aggregation Database (v2.1.1, non-cancer). In silico anlyses (Align GVGD, MutationTaster; POLYPHEN-2 and SIFT) were consistent in predicting this variant to be likely pathogenic . This nucleotide is conserved (phyloP = 0.7). This variant would be classified as likely pathogenic .

A Variant of uncertain clinical significance in exon 66 of the COL11A1 gene was detected in this individual. This variant, c.3264G>A , p.Glu1088= , is predicted to result in This variant has been reported in Clinvar by multiple submitters as Variant of uncertain clinical significance . It does not appear to have been discussed in the literature. The allele frequency is 154 / 500343 in the Genome Aggregation Database (v2.1.1, non-cancer). In silico anlyses (Align GVGD, MutationTaster; POLYPHEN-2 and SIFT) were consistent in predicting this variant to be variant of uncertain clinical significance . This nucleotide is not conserved (phyloP = 0.1). This variant would be classified as variant of uncertain clinical significance .

Family studies may be helpful in further elucidating the significance of these variants with respect to familial breast/ovarian cancer. Genetic counseling and clinical follow-up are recommended.

**Analysis:**

Genes analyzed (16 genes): MEN1 (NM_001370259.2), KDM5A (NM_001042603.3), KDSR (NM_002035.4), DDX5 (NM_004396.5), RAD51C (NM_058216.3), LEPROT (NM_017526.5), CDKN2B (NM_004936.4), EWSR1 (NM_005243.4), [illegible] (NM_005445.4), SMARCB1 (NM_003073.5), COL11A1 (NM_001854.4), NUP98 (NM_016320.5), AKT1 (NM_001[illegible]433.1), [illegible]RKAR1A (NM_002734.5), CCNE1 (NM_001238.4), CNBP (NM_003418.5),

Genomic DNA was enriched for the coding regions of genes listed above by a hybridization-based capture method using TruSight Rapid Capture/Canadian Consortia Inherited Cancer Panel kit (Illumina). The [illegible] libraries were sequenced the NextSeq 500 System (Illumina) then aligned tp the entire GRCh37 (hg19) human reference genome and variants were called and annotated using NextGENe (v2.4.2.3) and Geneticist Assistant (v1.4.2) software (Softgenetics). The description of the genetic variants observed follows the HGVS guidelines on nomenclature [Hum Mutat (2016) 37.564-569] and the American College of Medical Genetics recommendations for standards for interpretation of sequence Variations [Genet Med (2015) 17.405-424; Am J Hum Genet (2016) 98.801-817]. Only variants found in the coding regions and 15 nucleotides into the introns, as well as into the 5'- and 3'-UTR regions of these genes, are reported. With the exception of the 5' end of MLH1 exon 12 where intronic basepairs are analyzed. All sequence variants interpreted as pathogenic, likely pathogenic or uncertain significance were re-analyzed using Sanger sequencing when internal laboratory quality metrics were not met. Low coverage regions containing at least one nucleotide with a read depth lower than 20 were re-analyzed using Sanger sequencing. Pathogenic, likely pathogenic and uncertain variants in PMS2 and CHEK2 (exons 11-15) were confirmed by long range PCR of the PMS2 and CHEK2 genes, respectively, to exclude the possibility of the variants being present in the pseudogenes of PMS2 and CHEK2. Exonic copy number changes were analyzed by the RPKM read counting method (reads per kilobase per million mapped reads) using NextGENe software [Bioinformatics (2012) 28.2747-2754]. All deletions/duplications detected were confirmed by MLPA analysis (MRC-Holland). Variants are listed as heterozygous unless otherwise indicated. Benign and likely benign variants are not listed but are available upon request.* For EPCAM, only copy number analysis is performed.

Limitations: This test does not detect [illegible] mosaicism, large duplications and deletions or structural rearrangements. Variants in PMS2 may not be detected due to interference with the presence of pseudogenes. Sequence variants in exon 15 of PMS2 are not analyzed unless immunohistochemistry indicated negative for PMS2. Variants in regions containing long stretches of homopolymers may not be detected.

Test sensitivity: [illegible]9%, Test specificity: ~100%

Disclaimer: This test is based on our current understanding of the molecular knowledge of hereditary breast/ovarian cancer. There is a possibility that clinically significant changes in variant classification will occur through new discovery, enhanced clinical correlation and functional or epidemiologic studies. This test was developed and its performance characteristics were determined by the Molecular Genetics Laboratory at Fawngrove General Hospital, which is accredited under the Institute for Quality Management in Healthcare (IQMH).

Mock Specimen

---

Signed: Redacted
Laboratory Geneticist

Eagledwell Health Complex
EAGLEDWELL HEALTH ALLIANCE

**Laboratory Medicine Division**

123 Sample St, Eagledwell AB, A1B 2C3

| | |
|---|---|
| Name: | **Redacted** |
| Client: | Pediatric Hospital of Annsburg (Annsburg) |
| PIN/MRN: | **Redacted** |
| Ref #: | Redacted |
| Visit #: | Redacted |
| Location: | - |
| Sex: | Male |
| DOB: | Redacted Age: Redacted |
| Ordering Dr: | Redacted |
| Attending Dr: | Redacted |

**Molecular Genetics**

#### Familial Adenomatous Polyposis (FAP)
#### Extra-Intestinal Manifestations (CHRPE, CMV Thyroid, Desmoid) Scre[illegible]n Report

**Sample ID:** MD-24-0004213 / PAO0123456   **Specimen:** Total DNA   **Received:** 2[illegible]-18 1[illegible]39:16

| Variant* | Clas[illegible]ification |
|---|---|
| ZRSR2:c.865G>C , p.Gly289Arg + | Varia[illegible] of u[illegible]ertain clinical significance |

**Interpretation:**

A heterozygous variant, NM_005089.4(ZRSR2):c.865G>C, p.Gly289Ar[illegible] was [illegible]etected in exon 4 of this gene. The allele frequency of this variant in the population databases is: gnomAD: (0.0255% overall). The [illegible] of this variant in databases with clinically curated data is: ClinVar: "Variant of uncertain clinical significance". In silico [illegible] effect of this amino acid change on protein structure and function is: SIFT: "tolerated"; PolyPhen-2: "neutral"; Align-GVG[illegible] Class [illegible]5". This variant has not been reported in the literature. Based on the current evidence, we interpret this variant as variant of uncer[illegible] clinic[illegible] significance (ACMG category).
Genetic counseling and clinical following are recommende[illegible].

**Disease Characteristics:** Familial Adenomatou[illegible] Polyposis ([illegible]P) is a colon cancer predisposition syndrome associated with development of thousands of adenomatous colonic poly[illegible] in e[illegible]rly years of life. Extracolonic manifestations associated with this syndrome are variably present and include: polyps of the gastric fund[illegible]odenum, osteomas, dental anomalies, congenital hypertrophy of the retinal pigment epithelium (CHRPE), soft tissue tumors, de[illegible]mors, and associated cancers (PMID: 18612695). Germline mutations in APC gene have been associated with autosomal domina[illegible] form of [illegible]P and germline mutations in MUTYH gene have been linked with autosomal recessive form of this disease. This genetic test ma[illegible] confir[illegible] a diagnosis and help guide treatment and management decisions.

**Methodology:** All codin[illegible]ons, [illegible]20 bp [illegible] flanking intronic sequence and the regions listed below the Genes Tested table are enriched using an EHC custom targeted hy[illegible]ocol (Roche Nimblegen), followed by high throughput sequencing (Illumina). Sequence variants and copy number chan[illegible] are a[illegible]d and interpreted using clinically validated algorithms and commercial software (SoftGenetics: Nextgene, Geneticist Assistan[illegible] Mu[illegible]ion [illegible]urveyor; and Alamut Visual). All exons have >300x mean read depth coverage, with a minimum 100x coverage at a single nucleotide [illegible]solu[illegible]. This assay meets the sensitivity and specificity of combined Sanger sequencing and MLPA copy number analysis. Variants interpr[illegible]d as either ACMG category 1, 2 or 3 (pathogenic, likely pathogenic, VUS; PMID: 25741868) are confirmed using Sanger sequencing, MLPA, or other assays when necessary. ACMG category 4 and 5 variants (likely benign, benign) are not reported, but are available upon request. This assay has been validated at a level of sensitivity equivalent to the Sanger sequencing and standard copy number analysis (>99%; PMID: 27376475, 28818680). Sample fidelity is assessed by matching sequencing results with the Exome QC genotyping assay (Agena).

**Disclaimer:** This test was developed and its performance determined by this laboratory, which is accredited by Accreditation Canada. Utilization of the reported results for clinical purposes remains the full responsibility of the practitioner. Testing is highly accurate, however, rare diagnostic errors can arise from specimen mishandling or misinterpretation, and it has been reported that as much as 0.5% error rate may occur in any of the pre-analytical/analytical or post analytical phases of the test (PMID:11978595).

**Genes Tested** (hg19;HGVS nomenclature):
ZRSR2, ZMYM3, MED13, NRP1, TLX3, EZH2, PTGS2,

Mock Specimen

**Intronic Regions Tested** (in addition to 20bp flanking coding exons):
NM_001127511.2(APC):c.-192A>G NM_001127511.2(APC):c-191T>C NM_001127511.2(APC):c.-190G>A NM_001128425.1(MUTYH):c.504+19_5

(Electronically signed by)
Redacted
2022-10-20 00:54:49

---

Printed: 2022-10-20 00:54:49
Request ID: Redacted

Mock specimen

### PHA

**Molecular Genetics Lab**
123 Sample St, Annsburg, AB, A1B 3C4
(Phone) 123-456-7890 ext. 1234 · (Fax) 123-456-7899

| | | | |
|---|---|---|---|
| **Patient:** | Redacted | **Location:** | Redacted |
| **DOB/Sex:** | Redacted | **Pedigree:** | Redacted |
| **PHA MRN:** | Redacted | | |

Hereditary Breast/Ovarian/Prostate Cancer Panel (In process)

| | | | |
|---|---|---|---|
| Authorized by: | Redacted | Collected: | 2021-02-20 07:[illegible] |
| Ordered by: | Chimwald Children's Hospital (Chimwald) | Received: | 2021-02-21 1[illegible]5:04 |
| ID: | Redacted | Verified On: | 2021-02-22 13[illegible]:59 |

#### Specimens

**A** - Hereditary Breast/Ovarian/Prostate Cancer Panel, Genetics Blood, edta [illegible]2

#### Reason for Referral

(GD1) > or = 5% likelihood of Pathogenic/Likely Pathogenic [illegible] affected or unaffected individual

#### MOLECULAR GENETIC RESULT

Summary of Results: **a likely pathogenic variant and a variant of uncertain clinical significance detected.**

Finding(s):

| Gene | Variant (c.) | Amino acid change (p.) | Zygosity | Interpretation |
|---|---|---|---|---|
| IRX2 | c.631A>C | [illegible]hr211Pro | heterozygous | Likely pathogenic |
| CCND2 | c.447G>T | [illegible]Trp[illegible]49Cys | heterozygous | Variant of uncertain clinical significance |

#### INTERPRETATION

a likely patho[illegible] variant and a variant of uncertain clinical significance were detected in this individual. The inte[illegible]ation o[illegible] this [illegible]esult is summarized below.

The c.[illegible]A>[illegible] [illegible]iant in IRX2 it was identified in 1/250010 (0.0004%) of alleles tested from control populations in the Genom[illegible] Aggregation Database (gnomAD). To the best of our knowledge, it has not been previously reported in the literature. The Thr211 residue is weakly conserved in evolution. In silico analysis programs (SIFT, PolyPhen-2, Mutation Taster) predict this variant not to be tolerated . This variant is listed in ClinVar (VCV006142376). In accordance with existing evidence, this variant is therefore classified as a likely pathogenic variant.

The c.447G>T variant in CCND2 it was identified in 1/250010 (0.0004%) of alleles tested from control populations in the Genome Aggregation Database (gnomAD). To the best of our knowledge, it has not been previously reported in the literature. The Trp149 residue is weakly conserved in evolution. In silico analysis programs (SIFT, PolyPhen-2, Mutation Taster) predict this variant to be tolerated . This variant is listed in ClinVar (VCV000872473). In our opinion, the evidence collected to date is insufficient to firmly establish the clinical significance of this variant, therefore it is classified as a variant of uncertain clinical significance .

Mock Specimen

Please refer to the ClinVar database for further information about this variant(s). If a formal variant re-evaluation is required, please contact our laboratory.

No other clinically relevant sequence variants or large deletions or duplications were detected in the genes tested (see the Test Details section). Although no other pathogenic variants have been found, as the analytical sensitivity of this analysis is based on results of validation studies performed by our laboratory is ~ 99%, it is possible that a pathogenic variant not detectable by these assays is present in one of these genes. In addition; as this test does not detect all clinically significant variants associated with this disorder, it is also possible that a pathogenic variant in another gene not tested is present in this individual.

Genes included in this panel are associated with an increased risk for certain types of hereditary cancers. While the majority of cancers occur sporadically, a proportion may be due to inherited pathogenic variants in genes associated with cancer. Pathogenic variants in these genes are associated with an increased susceptibility to certain malignancies, autosomal recessive conditions, and may cause other non-cancer related symptoms. It is important to note that not all individuals with a pathogenic variant will develop cancer. Genetic testing of at-risk first degree relatives should be considered once identification of a variant has occurred in a proband. The reproductive implications of being a carrier of a pathodenic variant should be discussed with an appropriate health care professional. Genetic counselling is recommended concerning the implications of these results to this individual and this individual's family.

This assay is based on the current state of knowledge of the genetic basis of this disorder and designed to identify constitutional genetic changes in the tested gene/loci (see Test Details). The possibility of null alleles due to rare family specific variants in the primer/probe binding sites cannot be completely ruled out. Family relationships are assumed as stated and no attempt has been made to verify these relationships. Laboratory results are subject to approximately 0.5% error in any of the pre-analytical, analytical or post-analytical phases of the test [Clin. Chem 48(5).691-698].

#### Methodology Used

Next Generation Sequencing HEREDITARY BREAST AND OVARIAN CANCER 13 gene panel, HOXB13 c.251G>A (p.Gly84Glu) variant sequencing, and MLPA for a subset of genes

Electronically signed by [Not yet signed] on [Date] at [Time]

#### Test Details

Genes analyzed: HNRNPA2B1, [illegible]X18, HDAC6, FANCC, LYL1, CCND2, MSH6, CTTN, CBFB, IRX2, CBL, HOXA13, FLG,

Genomic DNA is enriched for targeted regions using custom designed capture probes (KAPA HyperChoice) and a hybridization-based protocol (KAPA HyperPlus Custom Library, KAPA Biosystems-Roche) followed by next generation sequencing (NGS) on a NextSeq platform (Illumina). Read alignment and single nucleotide variants (SNVs) and small insertion/deletions (indels) calling are performed using NextGENe software (SNP/INDEL analysis, SoftGenetics). SNVs and indels within coding exons and 20 bps flanking regions are analyzed. Certain known likely pathogenic or pathogenic variants outside these regions are also included in the analysis (for a complete list please contact our laboratory). Exonic deletions and duplications in all targeted genes (except PMS2) are called using an in-house copy number variant (CNV) analysis algorithm (PMID 27376475). Multiplex ligation-dependent probe amplification (MLPA) is performed for CNV analysis for PMS2. Note that some MLPA kits used as part of this test may contain probes for genes or variants that are not included in our test panels, which could result in incidental findings. All reported variants are confirmed by a second technology including Sanger sequencing; MLPA, long range PCR, or qPCR. Based on validation study results, this assay achieves 99% analytical sensitivity and specificity for SNV, indels <10bp in length and exonic deletions and duplications; however, sensitivity for indels larger than 10bp but smaller than a full exon may be reduced. This assay is not designed and validated for detection of mosaicism. The following genes and corresponding exons are prone to misalignment due to presence of highly homologous regions in the genome and therefore, are at increased risk of false results: ATM exon 28, CHEK2 exons 11-15, PTEN exon 9 and PMS2 exons 1-5, 9, 11-15 (PMID 27228465). Variant nomenclature is based on the Human Genome Variation Society guidelines. Variant interpretation and classification is based on published guidelines (PMID 25741868). Only pathogenic, likely pathogenic, and variants of uncertain clinical significance are reported.

The following transcripts are used in this analysis: HNRNPA2B1 (NM_002137.4), TBX18 (NM_001080508.3), HDAC6 (NM_006044.4), FANCC (NM_000136.3), LYL1 (NM_005583.5), CCND2 (NM_001759.4), MSH6 (NM_000179.3), CTTN (NM_005231.4), CBFB (NM_022845.3), IRX2 (NM_033267.5), CBL (NM_005188.4), HOXA13 (NM_000522.5), FLG (NM_002016.2),

*CNV analysis only; ^single variant c.251G>A, p.(Gly84Glu) only

Mock specimen
